# Heterozygous variants in *PLCG1* affect hearing, vision, cardiac, and immune function

**DOI:** 10.1101/2024.01.08.23300523

**Authors:** Mengqi Ma, Yiming Zheng, Mingxi Deng, Shenzhao Lu, Xueyang Pan, Xi Luo, Michelle Etoundi, David Li-Kroeger, Kim C. Worley, Lindsay C. Burrage, Lauren S. Blieden, Aimee Allworth, Wei-Liang Chen, Giuseppe Merla, Barbara Mandriani, Catherine E Otten, Pierre Blanc, Jill A. Rosenfeld, Debdeep Dutta, Shinya Yamamoto, Michael F. Wangler, Undiagnosed Diseases Network, Ian A. Glass, Jingheng Chen, Elizabeth Blue, Paolo Prontera, Jeremie Rosain, Sandrine Marlin, Seema R. Lalani, Hugo J. Bellen

## Abstract

Phospholipase C isozymes (PLCs) hydrolyze phosphatidylinositol 4,5-bisphosphate (PIP_2_) into inositol 1,4,5-trisphosphate (IP_3_) and diacylglycerol (DAG), important signaling molecules involved in many cellular processes including Ca^2+^ release from the endoplasmic reticulum (ER). *PLCG1* encodes the PLCγ1 isozyme that is broadly expressed. Hyperactive somatic mutations of *PLCG1* are observed in multiple cancers, but only one germline variant has been reported. Here we describe seven individuals with heterozygous missense variants in *PLCG1* [p.(Asp1019Gly), p.(His380Arg), p.(Asp1165Gly), and p.(Leu597Phe)] who present with hearing impairment (5/7), ocular pathology (4/7), cardiac septal defects (3/6), and various immunological issues (5/7). To model these variants *in vivo*, we generated the analogous variants in the *Drosophila* ortholog, *small wing* (*sl*). We created a null allele *sl^T2A^* and assessed its expression pattern. *sl* is broadly expressed, including wing discs, eye discs, and a subset of neurons and glia. *sl^T2A^*mutant flies exhibit wing size reductions, ectopic wing veins, and supernumerary photoreceptors. We document that mutant flies also exhibit a reduced lifespan and age-dependent locomotor defects. Expressing wild-type *sl* in *sl^T2A^* mutant flies rescues the loss-of-function phenotypes, whereas the variants increase lethality. Ectopic expression of an established hyperactive *PLCG1* variant, p.(Asp1165His) in the wing pouch causes elevated Ca^2+^ activity and severe wing phenotypes. These phenotypes are also observed when the p.(Asp1019Gly) or p.(Asp1165Gly) variants are overexpressed in the wing pouch, arguing that these are gain-of-function variants. However, the wing phenotypes associated with p.(His380Arg) or p.(Leu597Phe) overexpression are either mild or only partially penetrant. Our data suggest that the heterozygous missense variants reported here affect protein function differentially and contribute to the clinical features observed in the affected individuals.

## Introduction

The inositol lipid-specific phospholipase C (PLC) isozymes are key signaling proteins that play critical roles in transducing signals from hormones, growth factors, neurotransmitters, and many extracellular stimuli (Berridge and Irvine 1984; Exton 1996; Balla 2013). The PLCs selectively hydrolyze phosphatidylinositol 4,5-bisphosphate (PIP_2_) into inositol 1,4,5-trisphosphate (IP_3_) and diacylglycerol (DAG) (Nishizuka 1984; Majerus et al. 1986). PIP2 functions as a membrane anchor for numerous proteins and affects membrane dynamics and ion transport (Hilgemann et al. 2001; Hilgemann 2007; Suh and Hille 2008). The two products, IP_3_ and DAG, are important intracellular second messengers involved in Ca^2+^ signaling regulation and protein kinase C signaling activation, respectively (Nishizuka 1992; Berridge 1993). Hence, PLC orchestrates diverse cellular processes and behaviors, including cell growth, differentiation, migration, and cell death (Yang et al. 2012; Cocco et al. 2015; Gomes et al. 2021; Asano et al. 2022). There are at least thirteen PLC isozymes grouped in 6 classes (β, δ, ε, γ, η, ζ) in mammals with similar enzymatic function, but each PLC has its own spectrum of activators, expression pattern, and subcellular distribution (Suh et al. 2008; Kadamur and Ross 2013; Katan and Cockcroft 2020).

*PLCG1* [MIM: 172420] encodes the PLCγ1 isozyme. PLCγ1 can be directly activated by receptor tyrosine kinases (RTKs) as well as cytosolic receptors coupled to tyrosine kinases (Gresset et al. 2012). Upon tyrosine phosphorylation, PLCγ1 undergoes conformational changes that release its autoinhibition upon which it associates with the plasma membrane to bind and hydrolyze its substrates (Gresset et al. 2010; Hajicek et al. 2019; Nosbisch et al. 2022). There is a second PLCγ isozyme, PLCγ2, encoded by *PLCG2* [MIM: 600220]. Although these two isozymes have similar protein structure and activation mechanism, they are differentially expressed and regulated, and play non-redundant roles (Homma et al. 1989; Regunathan et al. 2006). *PLCG2* is mostly expressed in cells of the hematopoietic system and mainly functions in immune response, causing human diseases associated with immune disorders (Yu et al. 2005; Ombrello et al. 2012; Zhou et al. 2012; Neves et al. 2018; Baysac et al. 2024). However, *PLCG1* is ubiquitously expressed and is enriched in the central nervous system (CNS) (Consortium 2015). *Plcg1* is essential in mice, and a null allele causes embryonic lethality with developmental defects in the vascular, neuronal, and immune system (Ji et al. 1997; Liao et al. 2002). *PLCG1* has emerged as a possible driver for cell proliferation, and increased expression levels of *PLCG1* have been observed in breast cancer, colon cancer, and squamous cell carcinoma (Arteaga et al. 1991; Noh et al. 1994; Park et al. 1994; Xie et al. 2010). Moreover, hyperactive somatic mutations of *PLCG1* have been observed in angiosarcomas and T cell leukemia/lymphomas (Behjati et al. 2014; Kunze et al. 2014; Vaque et al. 2014; Kataoka et al. 2015). However, the genotype-phenotype association of germline *PLCG1* variants has yet to be explored.

Here, we reported seven individuals carrying heterozygous variants in *PLCG1* (GenBank: NM_002660.3) who exhibit partially overlapping clinical features including hearing impairment (5/7), ocular pathology (4/7), cardiac defects (3/6), abnormal brain MRI findings (2/3), and immunological issues with diverse manifestations (5/7). Utilizing *Drosophila* to model the variants *in vivo*, we provide evidence that the missense *PLCG1* variants are toxic and affect protein function to varying degrees. We argue that these variants contribute to the clinical symptoms observed in the affected individuals.

## Results

### Individuals with heterozygous missense variants in *PLCG1* exhibit hearing impairment, cardiac defects, ocular pathology, and immune dysregulation

Seven individuals with heterozygous missense variants in *PLCG1* were recruited trough the Undiagnosed Diseases Network (UDN) (Splinter et al. 2018) (Individuals 1-2) and GeneMatcher (Sobreira et al. 2015) (Individuals 3-7). Individuals 1-3 are *de novo* cases from unrelated families: Individual 1 [c.3056A>G, p.(Asp1019Gly)], Individual 2 [c.1139A>G, p.(His380Arg)] and Individual 3 [c.3494A>G, p.(Asp1165Gly)]. Individuals 4-7 are from the same family, and all carry the *PLCG1* variant [c.1789C>T p.(Leu597Phe)]. The phenotypes of the individuals partially overlap but show a spectrum of clinical manifestations. **[*As per medRxiv policy, the whole and detailed case history for the probands have been removed. To obtain more detailed information, please contact the authors]***

### The missense *PLCG1* variants affect conserved protein domains and are predicted to be deleterious

*PLCG1* is predicted to be tolerant to loss-of-function alleles with a pLI score (Lek et al. 2016) of 0.16, suggesting that loss of one copy of the gene is unlikely to cause haploinsufficiency in humans, consistent with the presence of many protein truncating variants in gnomAD (Karczewski et al. 2020). However, the missense constraint Z score (Lek et al. 2016) of *PLCG1* is 3.69, suggesting that it is intolerant to missense variants. Consistently, all variants are located within regions or stretches depleted in missense variants according to scores such as regional missense constraint (RMC) (Chao et al. 2024) or missense tolerance ratio (MTR) (Sun et al. 2024). In addition, the prediction based on the DOMINO algorithm indicates that *PLCG1* variants are likely to have a dominant effect (Quinodoz et al. 2017). Several other *in-silico* pathogenicity predictions also suggest that these variants are likely to be pathogenic (Table S1) based on MARRVEL (Wang et al. 2017).

The four variants identified from the affected individuals map to different conserved domains of PLCγ1, and each variant affects an amino acid residue that is conserved from flies to humans (Figure 1A and 1B). The p.(Asp1019Gly) and p.(His380Arg) variants map to the catalytic core domains (X and Y regions, respectively), the p.(Asp1165Gly) variant is in the C-terminal C2 domain and the p.(Leu597Phe) variant is in the nSH2 domain. The latter is part of the PLCγ-specific regulatory array composed of a split PH domain (sPH), two Src homology 2 (nSH2 and cSH2) domains and a Src homology 3 (SH3) domain. PLCγ1 also contains other conserved domains including an N-terminal pleckstrin homology (PH) domain and four EF hand motifs.

**Figure 1.**
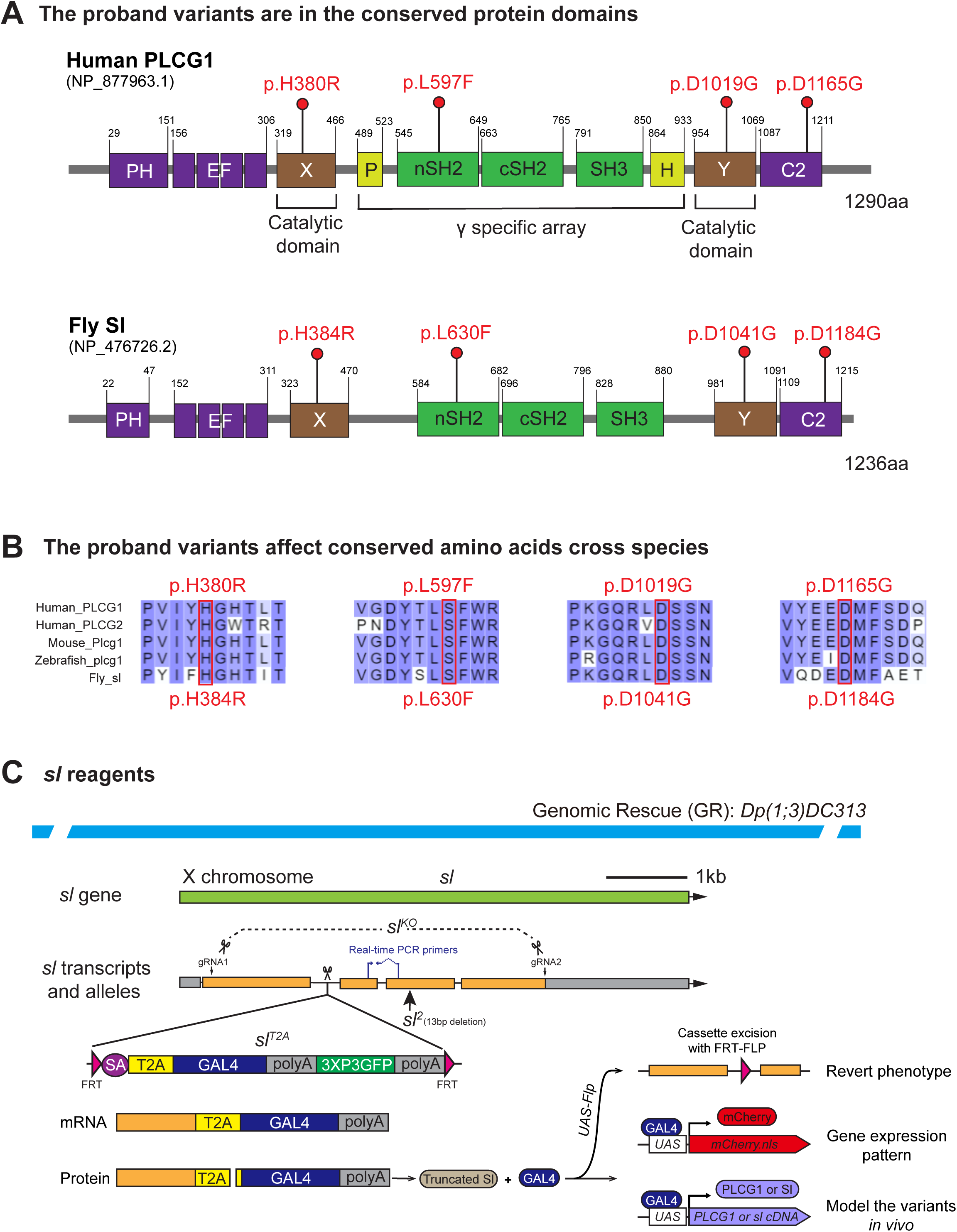
The *PLCG1* ortholog is *small wing* (*sl*) in *Drosophila*. (A) Schematic of human PLCG1 and fly Sl protein domains and positions of the variants identified in the affected individuals. Domain prediction is based on annotation from NCBI. (B) Alignment of protein domains near variants of PLCG1 and PLCG2 with PLCG1 from other species. The variants are marked with boxes. All the variants affect conserved amino acids (labeled in red). Isoforms for alignment: Human PLCG1 NP_877963.1; Human PLCG2 NP_002652.2; Mouse Plcg1 NP_067255.2; Zebrafish plcg1 NP_919388.1; Fly sl NP_476726.2. (C) Schematic of fly *sl* genomic span, transcript, alleles and the 92kb genomic rescue (GR) construct. Loss-of-function alleles of *sl* including *sl*^2^ (13bp deletion (Thackeray et al. 1998)), *sl^KO^* (CRISPR-mediated deletion of the gene span (Trivedi et al. 2020)), and *sl^T2A^*(T2A cassette inserted in the first intron, (Lee et al. 2018)) are indicated. The T2A cassette in *sl^T2A^* is flanked by FRT sites and can be excised by Flippase to revert loss-of-function phenotypes. GAL4 expression in *sl^T2A^* is driven by the endogenous *sl* promoter, allowing assessment of *sl* gene expression pattern with a *UAS-mCherry.nls* reporter line. This system also allows *in vivo* modeling of proband-associated variants by crossing with human *PLCG1* cDNAs or corresponding fly *sl* cDNAs. The primer pair used for real-time PCR is indicated.

### The *small wing* (*sl*) is the fly ortholog of human *PLCG1*

Flies have three genes encoding PLC isozymes (Table S2). Among them, *small wing* (*sl*) is predicted to be the ortholog of *PLCG1* with a DIOPT (DRSC Integrative Ortholog Prediction Tool) score of 17/18 (DIOPT version 9.0) (Hu et al. 2021). The encoded proteins share 39% identity and 57% similarity and are composed of similar conserved domains (Figure 1A). The *sl* gene is also predicted to be the ortholog of *PLCG2* with a DIOPT score of 12/18. These data suggest that *sl* corresponds to two human genes encoding the PLCγ isozymes. To obtain information about the nature of the *PLCG1* variants, we utilize *Drosophila* to model them *in vivo* using the binary GAL4 system (Brand and Perrimon 1993). We generated transgenic flies carrying the *UAS-human PLCG1* cDNAs for both the reference (*UAS-PLCG1^Reference^*) and the variants (*UAS-PLCG1^D1019G^*, *UAS-PLCG1^H380R^*, *UAS-PLCG1^D1165G^*, and *UAS-PLCG1^L597F^*). Given the high level of protein sequence homology and the conservation of the affected amino acids (Figure 1B), we also generated transgenic flies for the reference and analogous variants in the fly *sl* cDNA, namely *UAS-sl^WT^* and *UAS-sl^variants^* (*UAS-sl^D1041G^*, *UAS-sl^H384R^*, *UAS*-*sl^D1184G^*, and *UAS-sl^L630F^*).

In *Drosophila*, *sl* is on the X chromosome, and several alleles of *sl* have been isolated or previously generated, including *sl^2^*, *sl^KO^* and *sl^T2A^*(Figure 1C). *sl^2^* carries a 13bp deletion in the third exon that leads to a frameshift and early stop gain (Thackeray et al. 1998). *sl^2^* is a strong loss-of-function allele that causes small wing size, ectopic wing veins and extra R7 photoreceptors (Thackeray et al. 1998). *sl^KO^* was generated by CRISPR-mediated genomic editing that removes nearly the entire gene (Trivedi et al. 2020). *sl^T2A^* allele was generated by inserting an FRT-SA-T2A-GAL4-polyA-FRT cassette as an artificial exon into the first coding intron of *sl* (Figure 1C) (Diao et al. 2015; Lee et al. 2018). The polyA arrests transcription, and *sl^T2A^* is a strong loss-of-function allele (Figure S1A). The T2A viral sequence triggers ribosomal skipping and leads to the production of GAL4 proteins (Donnelly et al. 2001; Diao and White 2012) that are expressed in the proper spatial and temporal pattern of *sl*. This allows us to assess the expression pattern of *sl* by driving the expression of a *UAS-fluorescent protein* (Lee et al. 2018), or to assess the function of variants by expressing the human *UAS-reference/variant cDNAs* (Huang et al. 2022a; Huang et al. 2022b; Lu et al. 2022a; Lu et al. 2022b; Ma et al. 2023; Pan et al. 2023). In addition, the cassette is flanked by two FRT sites and can therefore be excised from the cells that express the gene in the presence of *UAS-Flippase* to revert the mutant phenotypes (Figure 1C) (Lee et al. 2018).

We first assessed the expression pattern of *sl* by driving *UAS-mCherry.nls* (an mCherry that localizes to nuclei) with *sl^T2A^*. *sl* is expressed in the 3^rd^ larval wing discs and eye discs (Figure 2A), consistent with the loss-of-function phenotypes observed in the wings and eyes (Thackeray et al. 1998). The expression pattern of *sl* in the wing discs is not homogenous. Higher expression levels are observed in the anterior compartment and along both the anterior/posterior and dorsal/ventral compartment boundaries (Figure 2A). The hemizygous *sl^T2A^/Y* male flies and the trans-heterozygous *sl^T2A^/sl^2^* or *sl^T2A^/sl^KO^*female flies show reduced wing size and ectopic wing veins (Figure 2B and Figure S1B), as well as additional photoreceptors in the eye (Figure 2C and Figure S1C). These phenotypes can be rescued by *UAS-Flippase* or by introducing a genomic rescue construct (*Dp(1;3)DC313* (Venken et al. 2010), Figure 1C) that covers the *sl* locus (Figure 2B and Figure 2C). These data show that all the observed phenotypes in *sl^T2A^* mutants can be attributed to the loss of *sl*.

**Figure 2.**
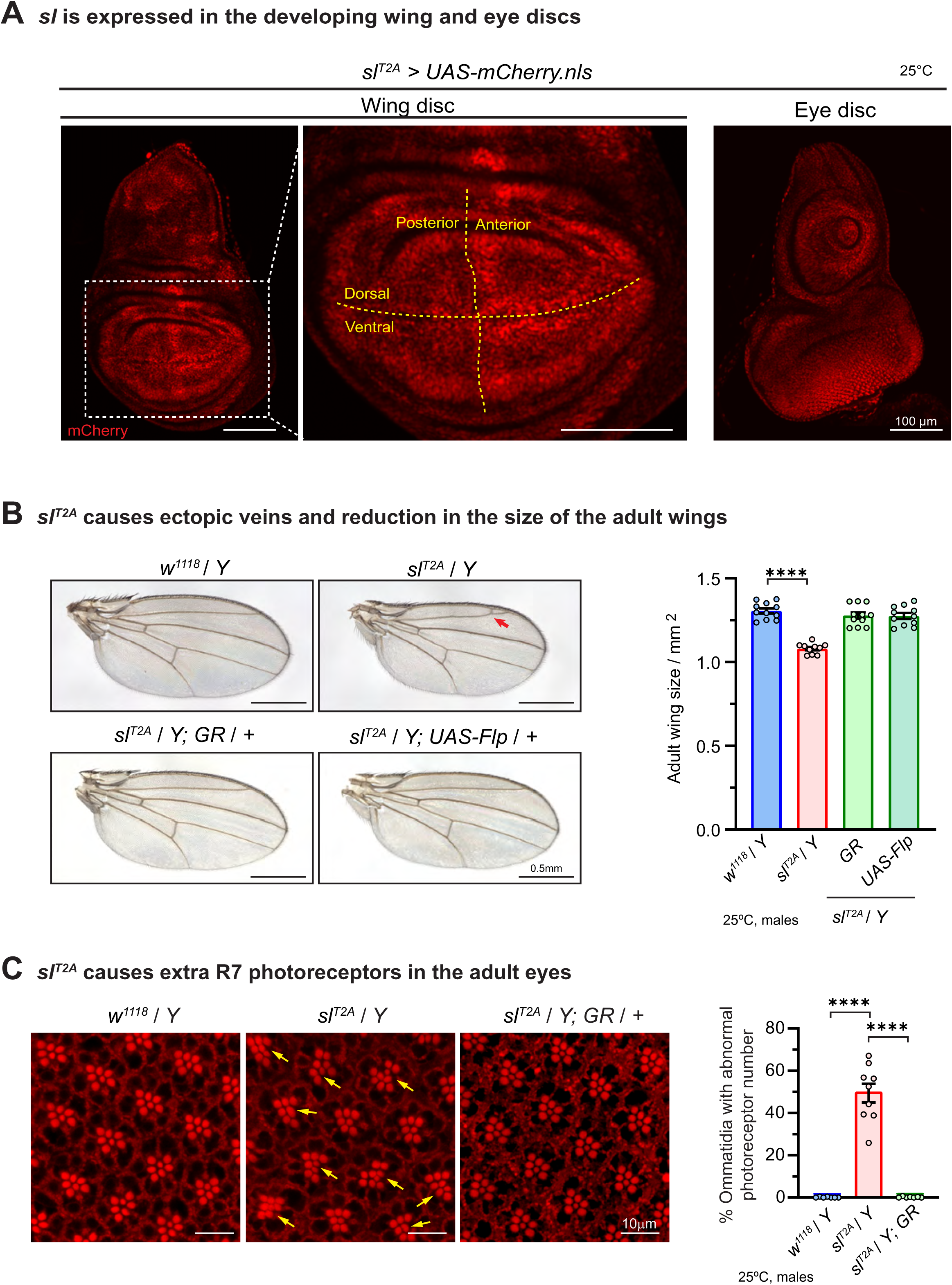
*sl^T2A^* is a loss-of-function allele that affect fly wing and eye development. (A) *sl* expression in wing and eye dises. Expression of *UAS-mcherry.nls* (red) was driven by *sl^T2A^*to label the nuclei of the cells that expressed *sl*. *sl* is expressed in the 3rd instar larval wing disc (left) and eye disc (right). Higher magnification image of the wing disc pouch region indicated by dashed rectangle is shown. The posterior/anterior and dorsal/ventral compartment boundaries are indicated by dashed lines in yellow. Scale bars, 100μm (B) *sl^T2A^* causes a wing size reduction and ectopic veins (arrowhead) in hemizygous mutant male flies. The wing phenotypes can be rescued by introduction of a genomic rescue (GR) construct or the expression of Flippase. Scale bars, 0.5mm. The quantification of adult wing size is shown in the right panel. Each dot represents the measurement of one adult wing sample. Unpaired t test, ∗∗∗∗p < 0.0001, mean ± SEM. (C) *sl^T2A^* causes extra photoreceptors (arrows) in the hemizygous mutant flies. The eye phenotype can be rescued by introduction of a genomic rescue (GR) construct. The photoreceptor rhabdomeres stain positive for phalloidin labeling F-actin. Scale bars, 10μm. The quantification is shown in the right panel. Each dot represents the measurement of one retina sample. Unpaired t test, ∗∗∗∗p < 0.0001, mean ± SEM.

### The *sl* gene is expressed in the fly CNS and loss of *sl* causes longevity and locomotion defects

Given that human *PLCG1* is highly expressed in the central nervous system (CNS) (Consortium 2015) and that the affected individuals present with neurologic phenotypes including hearing or vision deficits (Table 1), we investigated the expression pattern and the cell type specificity of *sl* in the CNS of flies. *sl* is expressed in the larval CNS as well as the adult brain, and co-staining with the pan-neuronal marker Elav (Robinow and White 1991) and glial marker Repo (Sepp et al. 2001) show that *sl* is expressed in many neurons and glia cells in the CNS (Figure 3A). We therefore assessed the longevity and climbing of *sl^T2A^* flies. Compared to the wild-type *w^1118^* flies, *sl^T2A^/Y* hemizygous mutant flies show a shortened lifespan and a progressively reduced climbing ability. These phenotypes can be rescued by expression of the wild-type *sl* cDNA(*sl^T2A^/Y; UAS-sl^WT^*) (Figure 3B).

**Figure 3.**
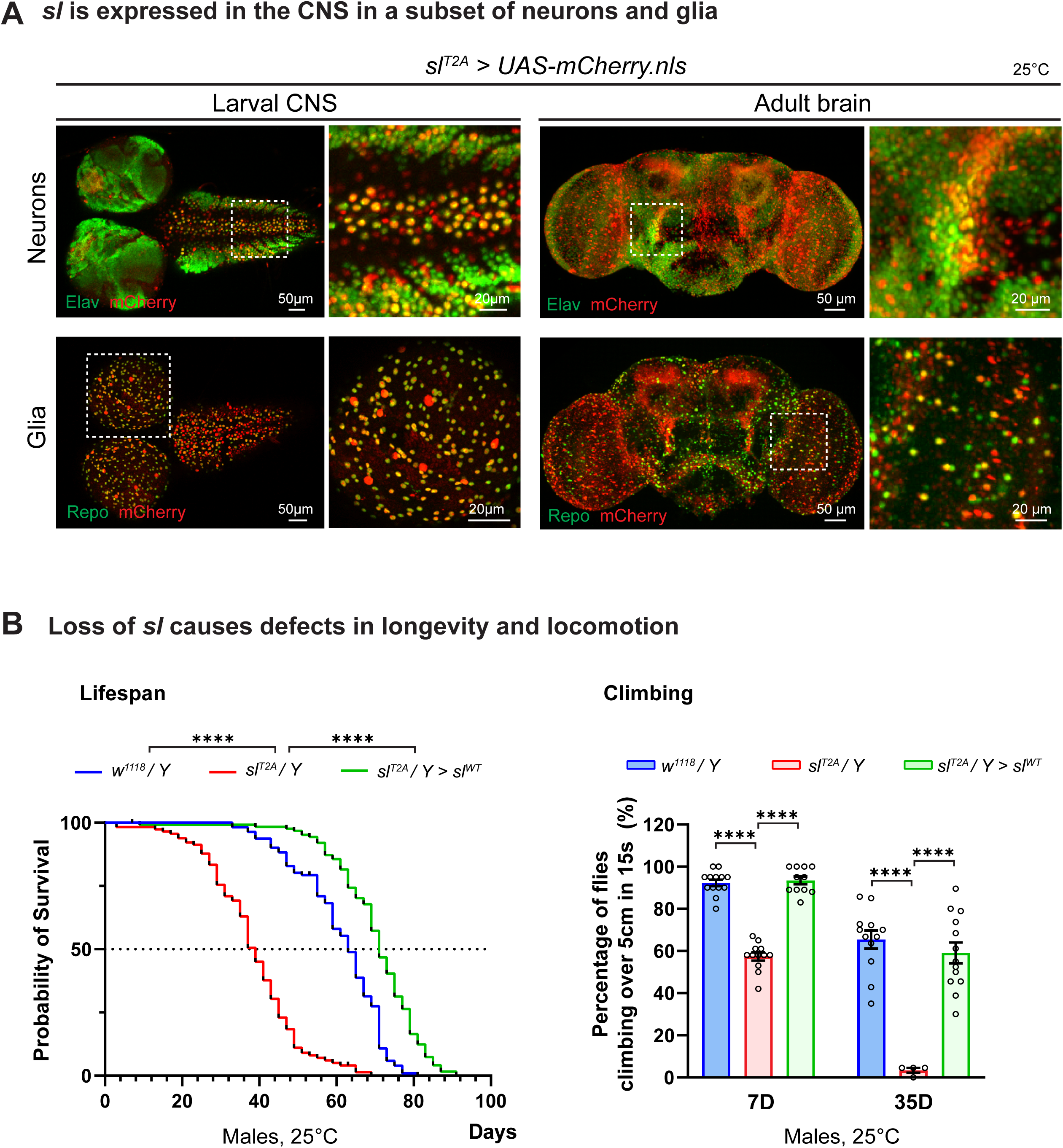
*sl* is expressed in a subset of neurons and glia in the CNS, and loss of *sl* causes behavioral defects. (A) Expression pattern of *sl* in the central nervous system observed by *sl^T2A^*-driven expression of *UAS-mCherry.nls* reporter (red). In either larval or adult brain, *sl* is expressed in a subset of fly neurons and glia, which were labeled by pan-neuronal marker Elav (green, upper panel) and pan-glia marker Repo (green, lower panel). Higher magnification images of the regions indicated by dashed rectangles are shown. Scale bars, 20μm in the magnified images, 50μm in other images. (B) Loss of *sl* causes defects in longevity and locomotion. *sl^T2A^* hemizygous flies have a shorter lifespan than *w^1118^* control flies. The median lifespan of *sl^T2A^* and *w^1118^* flies is 40 days and 62 days respectively. The shorter lifespan of *sl^T2A^* flies can be rescued by a UAS transgene that expresses the wild-type *sl* cDNA (*sl^WT^*). Fly locomotion was assessed by climbing assay (see methods). *sl^T2A^*flies at the age of 7 days show reduced locomotion and become almost immotile at the age of 35 days. The reduced locomotion ability in *sl^T2A^* flies can be fully rescued by *sl^WT^*. For lifespan assay, Longrank test, ****p<0.0001. For climbing assay, each dot represents measurement of one vial containing 18-22 flies for test. Unpaired t test, ****p<0.0001.

### Functional assays in flies indicate that the *PLCG1* variants are toxic

To assess the impact of the variants, we expressed the *sl* variant cDNAs in the *sl^T2A^/Y* hemizygous mutant males(*sl^T2A^/Y; UAS-sl^variants^*) and compared their rescue ability with the wild-type *sl*(*sl^T2A^/Y; UAS-sl^WT^*). As shown in Figure 4A (middle panel), the *sl^T2A^/Y* mutant flies (or the ones expressing a *UAS-Empty* control construct) have a slightly reduced eclosion rate, but expression of the *sl^WT^* cDNA fully rescues the percentage of eclosing progeny as measured by the Mendelian ratio. In contrast, expression of *sl^L630F^* (*sl^T2A^/Y; UAS-sl^L630F^*) reduced the percentage of hemizygous male progeny from the expected 25% to approximately 17%, while expression of *sl^H384R^* causes a severe reduction in the number of eclosing flies, with only a few escapers (*sl^T2A^/Y; UAS-sl^H384R^*). Expression of the *sl^D1041G^* or *sl^D1184G^* leads to 100% lethality. These data clearly indicate that these variants are toxic but at different levels.

**Figure 4.**
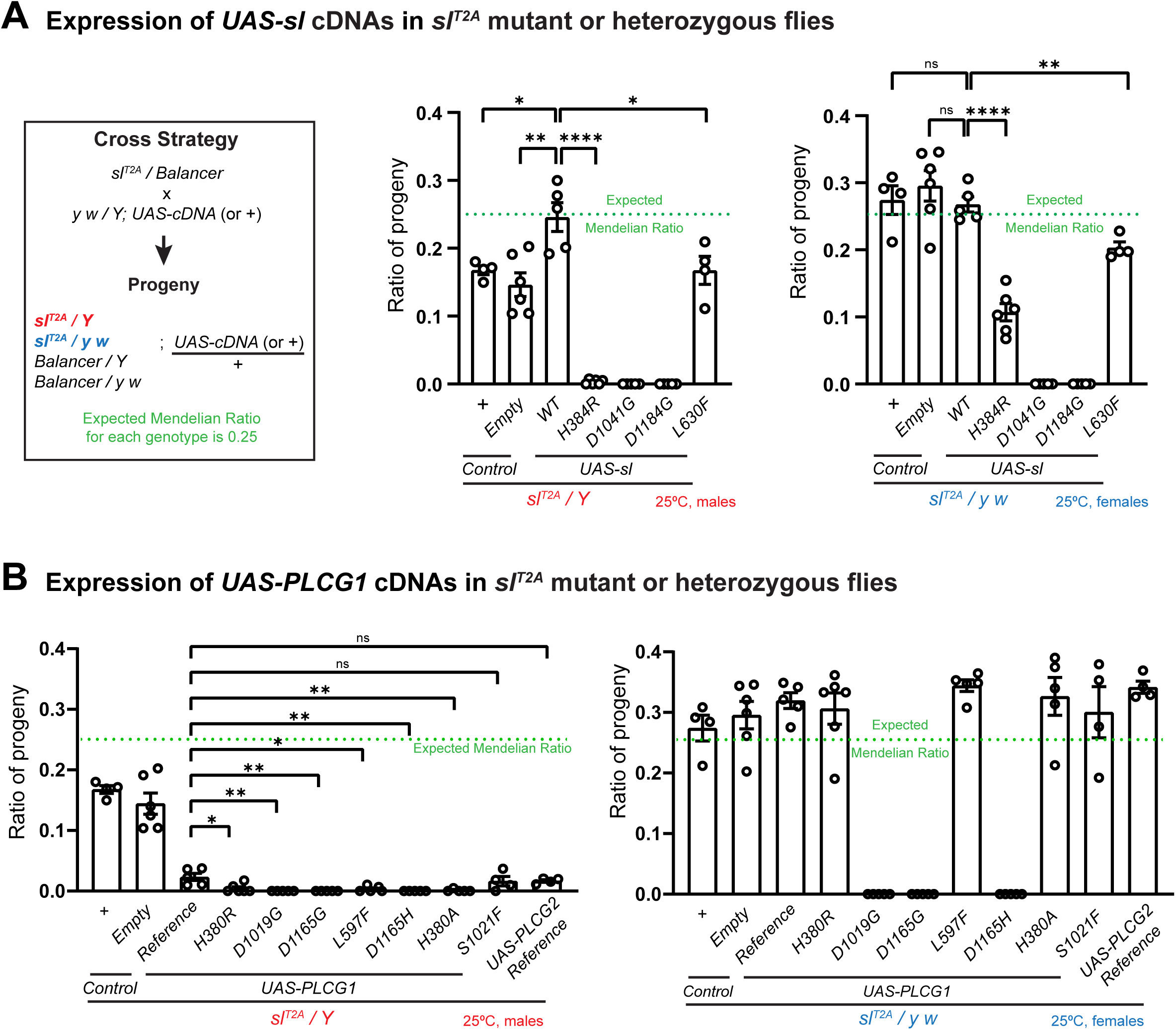
The human and corresponding fly variants are toxic when expressed in flies. (A) Summary of the viability associated with expression of *sl* cDNAs in *sl^T2A^* mutant or heterozygous flies. Cross strategy: heterozygous *sl^T2A^* female flies were crossed to male flies carrying *UAS-cDNAs* or control (*UAS-Empty*) constructs, or crossed to the *y w* males as an extra control. The percentages of hemizygous *sl^T2A^/Y* male progeny (red) or *sl^T2A^/yw* heterozygous female progeny (blue) that express different *UAS-cDNA* constructs were calculated. The expected Mendelian ratio is 0.25 (indicated by the green line in the graph). The fly analogue variants of the proband-associated variants were tested. Each dot represents one independent replicate. Unpaired t test, *p<0.05, **p < 0.01, ****p < 0.0001, ns: not significant, mean ± SEM. (B) Summary of the viability associated with the expression of *PLCG1* cDNAs in *sl^T2A^* mutant (red, males) or heterozygous (blue, females) flies. The same cross strategy and progeny ratio measurement described in (A) were applied. The proband-associated variants as well as three previously reported *PLCG1* variants were assessed. We also included the *PLCG2* reference cDNA. Each dot represents one independent replicate. Unpaired t test, *p<0.05, **p < 0.01, ns: not significant, mean ± SEM.

Since the *sl^T2A^/Y; UAS-cDNA* hemizygous males lack the endogenous *sl^+^,* we tested *sl^T2A^/yw; UAS-cDNA* heterozygous female flies that carry a copy of wild-type *sl^+^* while simultaneously expressing *UAS-cDNAs* driven by the *sl^T2A^* driver in the cells that endogenously express *sl* (Figure 4A, right panel). The eclosion rates of heterozygous female progeny expressing *sl* variants were significantly reduced compared to those expressing *sl^WT^*. Expression of *sl^H384R^* or *sl^L630F^* in the heterozygous progeny reduced the expected 25% proportion to approximately 10% and 20%, respectively, whereas expression of *sl^D1041G^* or *sl^D1184G^*resulted in complete lethality in heterozygous flies. These results suggest that the missense variants exert a dominant toxic effect. Additionally, we observed that the toxicity may have both developmental and acute effects in adults, with varying severity among the different variants (Figure S2), indicating that *sl* function is required in adult flies, implying that *PLCG1* variants may cause long-term deficits in affected individuals.

To compare the *sl* and *PLCG1* associated phenotypes, we conducted similar assays using human *PLCG1* cDNAs (Figure 4B). Expression of *PLCG1^Reference^* in the *sl^T2A^/Y* mutant flies (*sl^T2A^/Y; UAS-PLCG1^Reference^*) reduces viability by 80%, and expression of the other PLCγ coding gene, *PLCG2*, is also toxic and causes similar viability reduction compared to *PLCG1^Reference^*. This suggests that expression of human PLCγ genes is toxic in flies. This toxicity appears to be associated with expression level (Figure S3), and the survivals of *sl^T2A^/Y; UAS-PLCG1^Reference^* did not show rescue of the loss-of-function phenotypes in the wings or eyes (Figure S4). Expression of *PLCG1^H380R^*or *PLCG1^L597F^* in the *sl^T2A^/Y* mutant flies (*sl^T2A^/Y; UAS-PLCG1^H380R^*or *sl^T2A^/Y; UAS-PLCG1^L597F^*) leads to a significant but very modest increase in lethality when compared to *PLCG1^Reference^*, whereas expression of *PLCG1^D1019G^* or *PLCG1^D1165G^*results in 100% lethality (Figure 4B, left panel). When the reference and variants are assayed in the presence of a wild type copy of *sl^+^,* the heterozygous female progeny expressing the reference cDNA of *PLCG1* or *PLCG2* exhibited normal eclosion rate, as did the ones expressing *PLCG1^H380R^* or *PLCG1^L597F^*, suggesting that the presence of a wild-type copy of *sl^+^* combined with the reduced expression levels (typically 50% due to dosage compensation for the cells on X chromosome) mask some of the potential toxicity. However, expression of *PLCG1^D1019G^* or *PLCG1^D1165G^*still resulted in complete lethality in the females (Figure 4B, right panel). In summary, expression of the *PLCG1* variants and the corresponding fly *sl* variants exhibit greater toxicity than the reference or wild-type proteins with varying degrees of severity, suggesting that the variants are likely to be gain-of-function or neomorphic alleles. Among them, the *PLCG1^D1019G^* and *sl^D1041G^*, as well as *PLCG1^D1165G^* and *sl^D1184G^* are very strong toxic alleles whereas *PLCG1^H380R^, PLCG1^L597F^,* and their fly analogues are mild variants.

### The p.Asp1019Gly and p.Asp1165Gly variants are hyperactive

To assess whether the variants act as gain-of-function alleles that enhance the enzymatic activity of the PLCγ1 isozyme, we tested them using a Ca²D reporter assay. Since one of the products of the PLCγ1 isozyme, IPD, binds to receptors on the endoplasmic reticulum to trigger Ca²D release (Foskett et al. 2007), intracellular Ca²D levels can serve as a proxy of the PLCγ1 enzymatic activity. We expressed the *CaLexA* (calcium-dependent nuclear import of LexA) reporter (Masuyama et al. 2012) in the wing disc pouch region using a specific GAL4 driver (*nub-GAL4 > UAS-CaLexA.GFP*) while simultaneously expressing *UAS-PLCG1* cDNAs. We first assessed three control variants: *PLCG1^H380A^, PLCG1^D1165H^,* and *PLCG1^S1021F^*. Substitution of His380 with Ala (H380A) has been reported to suppress PIP_2_ hydrolysis and IP_3_ production (Smith et al. 1994; Wada et al. 2022), acting as an enzymatic-dead loss-of-function allele. On the other hand, the p.Asp1165His (D1165H) variant was previously identified as a strong gain-of-function somatic variant in adult T cell leukemia/lymphoma (Kataoka et al. 2015; Hajicek et al. 2019; Siraliev-Perez et al. 2022), and has been documented to cause a dramatic increase in phospholipase activity *in vitro* (Hajicek et al. 2019; Siraliev-Perez et al. 2022). The p.Ser1021Phe variant was reported recently in a *de novo* case and was characterized as a gain-of-function germline variant (Tao et al. 2023). As shown in Figure S5A, the GFP signal of the *CaLexA.GFP* reporter was low in wing discs expressing *PLCG1^H380A^*, whereas the signal was significantly enhanced in those expressing *PLCG1^D1165H^ or PLCG1^S1021F^*, showing that this is a robust assay for detecting increased enzymatic activity. We next tested the variants of the affected individuals. As shown in Figure 5A, expression of *PLCG1^Reference^* did not induce obvious GFP signals, suggesting that the protein is not enzymatically active, possibly because of autoinhibition. Similarly, expression of *PLCG1^H380R^* or *PLCG1^L597F^* did not significantly alter the GFP signal, suggesting that they are not constitutively active. However, expression of *PLCG1^D1019G^* or *PLCG1^D1165G^* markedly increased the GFP signal, similar to the *PLCG1^D1165H^*and *PLCG1^S1021F^* positive controls (Figure 5A and S4A). The same observations were made with the fly *sl* variants (Figure S5B). These results indicate that the *PLCG1^D1019G^* and *PLCG1^D1165G^* variants are hyperactive, whereas the *PLCG1^H380R^* and *PLCG1^L597F^* variants are not hyperactive based on this assay.

**Figure 5.**
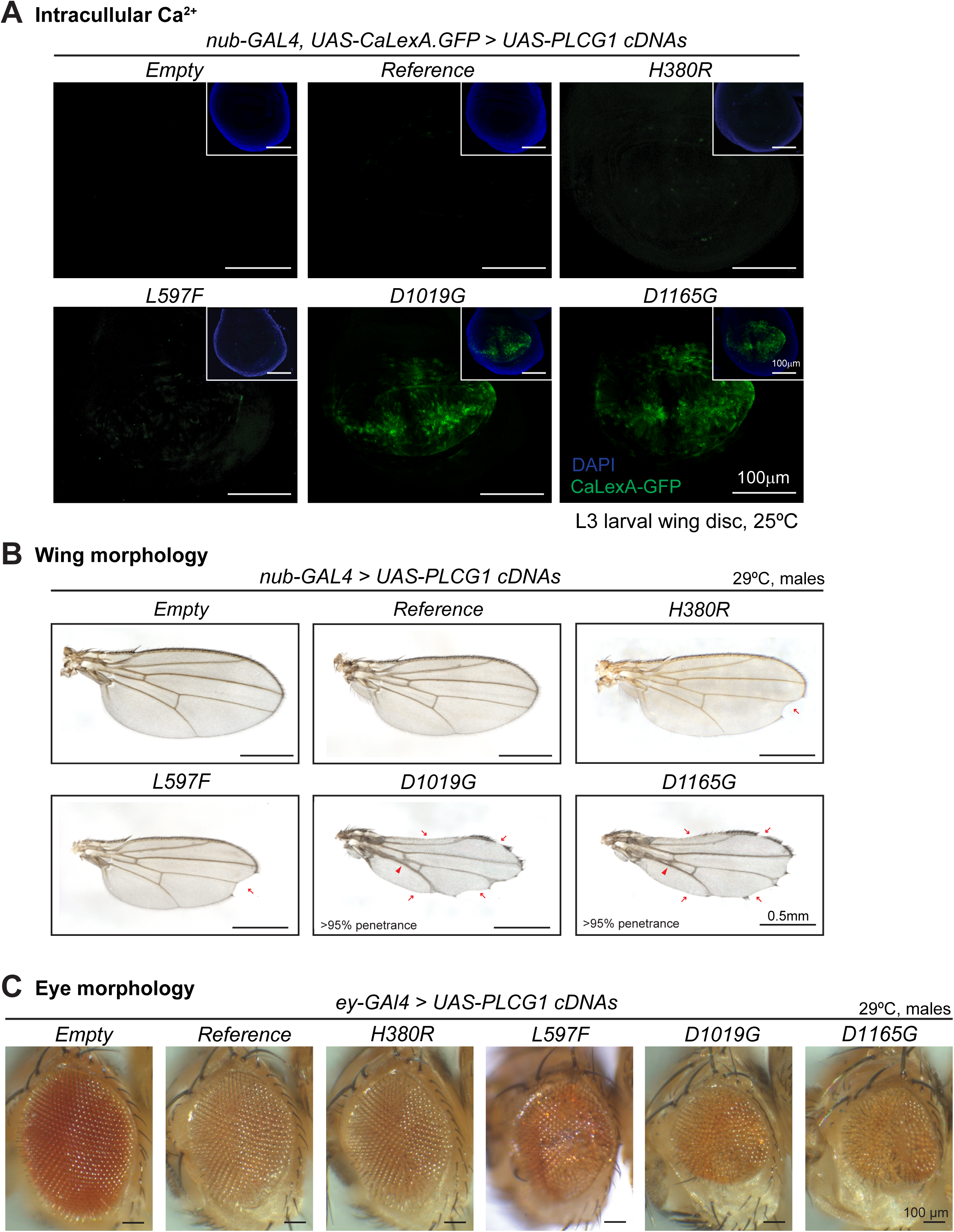
Ectopic expression of *PLCG1* variants causes variable phenotypes. (A) The Ca^2+^ reporter *CaLexA.GFP* was expressed in the wing disc pouch, simultaneously with the *PLCG1* cDNAs. Expression of *PLCG1^D1019G^* or *PLCG1^D1165G^*caused elevated CaLexA.GFP signal (green), indicating increased intracellular Ca^2+^ levels indicating that these variants are hyperactive. Nuclei were labeled with DAPI (blue). Scale bars, 100μm. (B) Representative images of the adult wing blades showing the morphological phenotypes caused by wing-specific expression of *PLCG1* cDNAs. Expression of *PLCG1^D1019G^* or *PLCG1^D1165G^* caused severe wing morphology defects including notched margin (arrows) and fused/thickened veins (arrowheads). Expression of *PLCG1^L597F^* exhibited partial penetrance (penetration ratio indicated). Expression of *PLCG1^H380R^* exhibited very mild phenotypes, comparable to *PLCG1^Reference^*. Scale bars, 0.5mm. (C) Representative images showing that eye-specific expression of *PLCG1^Reference^* or *PLCG1^H380R^* causes a ∼15% eye size reduction compared to the *UAS-Empty* control construct, and expression of *PLCG1^L597F^* further reduced eye size. Expression of *PLCG1^D1019G^* or *PLCG1^D1165G^*causes a severe size reduction by ∼30%. Each dot in the quantification graph represents the measurement of one adult eye. Unpaired t test, *p<0.05, ****p < 0.0001, ns: not significant, mean ± SEM. Scale bars, 100μm.

### The *PLCG1* variants affect size and morphology of wings and eyes

To further assess the impact of the *PLCG1* variants on normal development, we analyzed the morphology of the adult wings upon wing-specific expression of *PLCG1* or *sl* cDNAs (*nub-GAL4 > UAS-cDNAs*). Interestingly, ectopic expression of either *PLCG1^Reference^* or *sl^WT^* in the wing disc leads to a ∼10% reduction in adult wing size when compared to the *UAS-Empty* control (Figure S6A). This observation, together with the reduced wing size seen in the loss-of-function context (Figures 2B), suggests that both reduced and elevated levels of PLCγ1 can impair wing growth. This implies a dosage-dependent regulation on wing growth by the PLCγ1 isozymes, while the underlying mechanism is unknown. Additionally, as shown in Figure 5B and S6B, approximately 10% of the wings expressing *PLCG1^Reference^* exhibit notching along the wing margin, a phenotype not observed in wings expressing *sl^WT^*. Expression of *PLCG1^H380R^* or *PLCG1^L597F^* caused notched wings in approximately 18% and 23% of the flies, respectively (Figure S6B), whereas expression of *PLCG1^D1019G^* or *PLCG1^D1165G^* results in severe wing phenotypes characterized by notched wing margins, fused/thickened veins and reduced wing sizes with >95% penetrance (Figure 5B). Notably, expression of fly *sl^variants^*could lead to similar morphological defects as their corresponding human variants, arguing that these wing phenotypes are due to alterations of PLCG1 or Sl protein function (Figure 5B and S6C).

We also assessed the effect of expression of human *PLCG1* on eye development using the *eyeless-GAL4* (*ey-GAL4*). Expression of *PLCG1^Reference^*or *PLCG1^H380R^* in fly eyes leads to a mild reduction in eye size when compared to *UAS-Empty* control (Figure 5C and S6D). However, expression of *PLCG1^L597F^* results in rough eyes that are reduced in size whereas overexpression of *PLCG1^D1019G^* or *PLCG1^D1165G^*leads to a more severe eye phenotype (Figure 5C and S6D). In summary, the eye data are consistent with the wing data, showing that *PLCG1^D1019G^* and *PLCG1^D1165G^* are more toxic than *PLCG1^Reference^*. On the other hand, the toxicity of *PLCG1^H380R^* and *PLCG1^L597F^*are stronger than the *PLCG1^Reference^* but not as severe as *PLCG1^D1019G^* and *PLCG1^D1165G^*. Interestingly, the morphological defects in wings or eyes caused by ectopic expression of *PLCG1* cDNAs correlate with the expression level (Figure S6E), but do not directly correlate with the phospholipase enzymatic activity. For example, expression of *PLCG1^S1021F^* does not cause obvious morphological defects when compared to *PLCG1^Reference^* (Figure S6B and S6F), even though *PLCG1^S1021F^*is hyperactive and induces significantly elevated intracellular Ca^2+^ in the CaLexA reporter assay (Figure S5A).

## Discussion

Here we report seven individuals who carry heterozygous missense variants in *PLCG1* which encodes the phospholipase C γ1 isozyme. The individuals present with partially overlapping clinical features including hearing impairment, eye abnormality, heart defects and immune phenotypes. We show that the fly ortholog, *small wing* (*sl*), is widely expressed in wings and eyes, as well as in the central nervous system. Consistent with its expression pattern, we report that *sl* not only regulates wing and eye development, as previously documented, but also plays critical roles in the nervous system and affects locomotion and longevity. Furthermore, we assessed the function of the variants in the context of the human and fly cDNAs and show that their expression induces variable levels of toxicity when compared to the reference *PLCG1* or wild-type *sl*. Two of the variants are clearly hyperactive, and all the variants exhibit neomorphic effects (discussed in Supplemental Figure Notes). These observations show that the variants impair the normal function *in vivo* and suggest that they contribute to the symptoms observed in the affected individuals. Similarly to inborn error caused by the paralogous *PLCG2* (Baysac et al. 2024), germline variants in *PLCG1* can be pathogenic and dominant by different mechanisms.

### Structural analysis of the PLCG1 variants

Previously, studies based on biochemical assays and protein structures provided insights into how the variants studied here may affect the enzymatic activity of PLCγ1 (the protein structure of full-length rat Plcg1 is shown in Figure 6A). In its basal state, the PLCγ-specific regulatory array (sPH-nSH2-cSH2-SH3) forms autoinhibitory interfaces with the catalytic domains. Upon activation by the RTKs through binding with nSH2, PLCγ1 is phosphorylated, which induces the dissociation of the inhibitory cSH2 domain from the C2 domain. This triggers conformational rearrangements, allowing the enzyme to associate with the membrane and to expose the catalytic domains to allow hydrolysis of PIP2 (Gresset et al. 2010; Hajicek et al. 2019; Liu et al. 2020; Le Huray et al. 2022; Nosbisch et al. 2022). As shown in Figure 6A, the proband-associated variants map to conserved domains of the protein, either within the catalytic domains or at intramolecular and intermolecular interfaces. The p.Asp1019Gly and p.Asp1165Gly variants impact key residues involved in autoinhibition, leading to increased enzymatic activity. Specifically, the p.Asp1019Gly variant affects a conserved residue within the hydrophobic ridge of the Y box (Figure 6B), which is important for interaction with the sPH domain. This interaction is critical for the autoinhibition by blocking the membrane engagement of the catalytic core domain prior to enzymatic activation (Ellis et al. 1998; Hajicek et al. 2019). Notably, a substitution at the same position (Asp1019Lys, D1019K) has been demonstrated to enhance basal phospholipase activity *in vitro* (Hajicek et al. 2013), supporting its regulatory importance. Similarly, another hotspot somatic variant, p.Ser345Phe, located in the corresponding hydrophobic ridge within the X box, is also hyperactive (Vaque et al. 2014; Manso et al. 2015). On the other hand, the p.Asp1165Gly variant affects a residue situated within a loop of the C2 domain (Figure 6C). The Asp1165 residue plays a key role in stabilizing the interaction between the cSH2 domain and the C2 domain to maintain the autoinhibited state (DeBell et al. 2007). As mentioned above, the somatic variant p.Asp1165His leads to significantly elevated phospholipase activity *in vitro* (Liu et al. 2020; Siraliev-Perez et al. 2022), and results in severe phenotypes *in vivo* (Figure 4B, S5E and S5F). Molecular dynamics simulation data consistently indicate that autoinhibition is likely disrupted by the p.Asp1019Gly and p.Asp1165Gly variants (Figure S7A). In contrast, the p.His380Arg variant impacts the His380 residue within the X box, situated near a Ca^2+^ ion in the catalytic core (Figure 6D). His380 plays a role in coordination of the phosphate group at the 1-position of IP_3_ (Le Huray et al. 2022). While this residue may not be key to autoinhibition, it is important for the phospholipase activity. Substitution of His380 with Phe or Ala (H380F, H380A) have been reported to suppress PIP_2_ hydrolysis and IP_3_ production (Smith et al. 1994; Wada et al. 2022). Hence, substitution of the His380 with Arg in p.His380Arg variant may create a more basic environment, impacting the lipase activity. On the other hand, the p.Leu597Phe variant affects a residue within the nSH2 domain, which is part of the PLCγ-specific regulatory array (Figure 6E). The nSH2 domain mediates interactions with phosphorylated tyrosine residues on RTKs to initiate activation (Bae et al. 2009). Leu597 is located near the phosphotyrosine-binding pocket and this variant may therefore alter receptor specificity or induce novel protein interactions. Additionally, we utilized the DDMut platform (Zhou et al. 2023) to predict protein stability and folding of the variants, which are discussed in Figure S7B. In summary, our *in vivo* data are consistent with previous reports and *in silico* analyses, showing that the affected amino acids map to critical residues and strengthening the conclusion that the variants are pathogenic and likely impact the protein function through distinct mechanisms.

**Figure 6.**
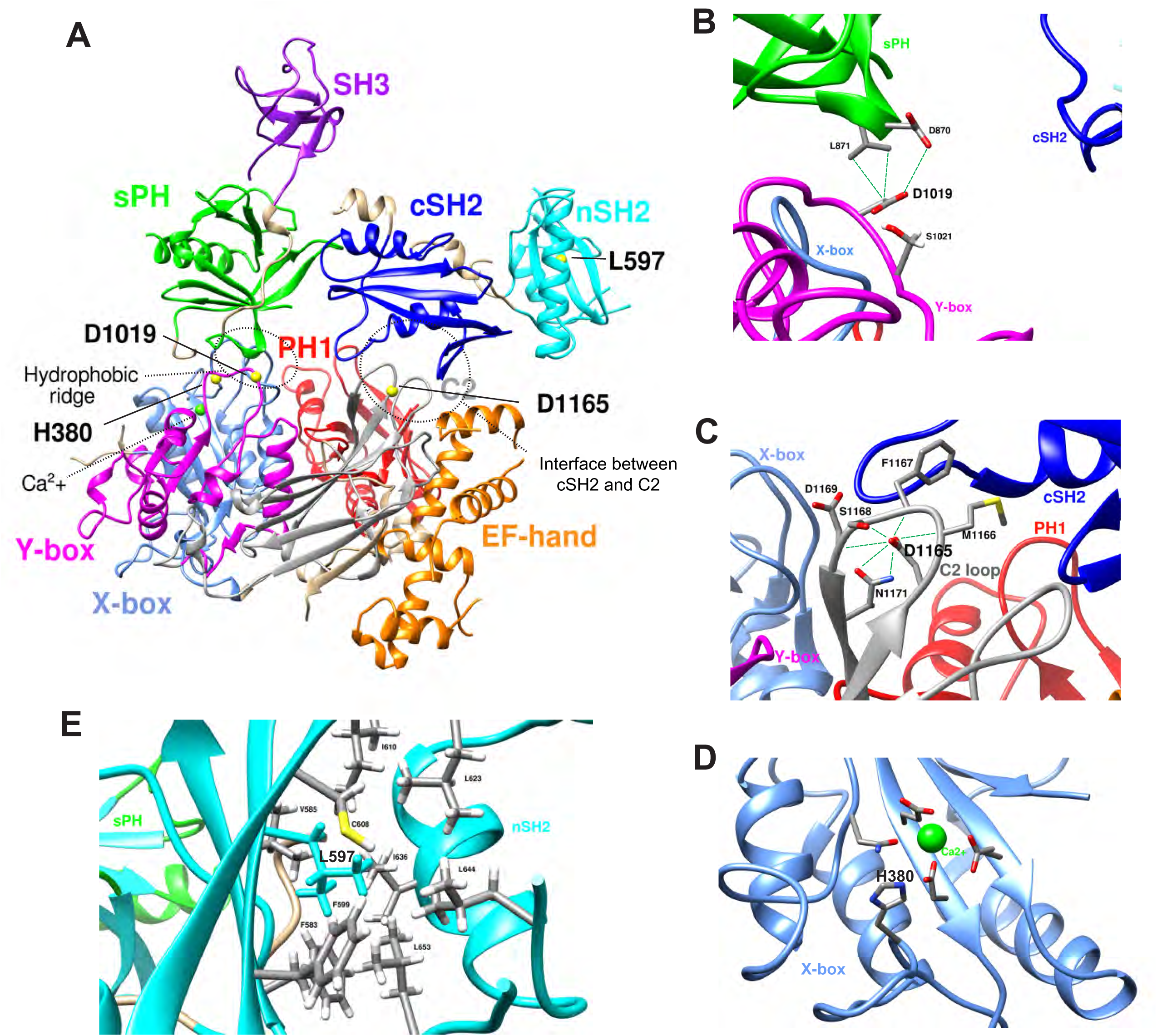
*PLCG1* variants affect important residues. (A) 3D structure of full-length rat Plcg1 (rat Plcg1 shares 97% amino acid identity with human PLCG1). The conserved protein domains are labeled with different colors. Two major intracellular interfaces are circled by dashed lines: 1-The hydrophobic ridge between the sPH domain and the catalytic core (X-box and Y-box); and 2-The interface between the cSH2 domain and the C2 domain. The four amino acids affected by the variants are shown as bolded black and indicated by yellow balls. (B) Enlarged views of the Asp1019 residue within the autoinhibition interface between sPH domain and the Y box. The potential interactions with nearby residues are indicated. (C) Enlarged view of the Asp1165 residue within the autoinhibition interface between the cSH2 domain and the C2 domain. The potential interactions with nearby residues are indicated. (D) Enlarged view of the His380 residue within the X-box catalytic domain, in proximity to the Ca^2+^ cofactor. (E) Enlarged view of the Leu597 and nearby residues in the nSH2 domain. Structural analysis was performed via UCSF Chimera (Pettersen et al. 2004)

### The *PLCG1* variants affect protein function to varying degrees and are associated with variable clinical manifestations

To better assess the genotype-phenotype relationship of the variants, we summarize the clinical features of affected individuals in Table 1, and the phenotypic effects observed in fly assays in Table S3. The p.(Asp1019Gly) (carried by Individual 1) and p.(Asp1165Gly) (carried by Individual 3) variants and their corresponding fly variants induce severe phenotypes across all assays performed. Individuals 1 and 3 share several obvious clinical features including hearing loss and heart septal defect. In contrast, the p.(His380Arg) and p.(Leu597Phe) variants cause mild or partially penetrant phenotypes across the different fly assays. Individual 2 who carries the p.(His380Arg) variant does not exhibit hearing impairment or heart defects observed in Individuals 1 and 3, but has eye malformations and neuroinflammation features that are shared with individuals 1 and 3, although the ocular and immunological defects manifest differently among individuals. Interestingly, individuals 4-7 are from the same family and all carry the p.(Leu597Phe) variant but also differ in their phenotypes, yet all share some clinical features with Individuals 1-3 (Table 1).

The heterogeneity in clinical manifestations may be influenced by additional genetic variants (see Table 1 legend) and environmental factors. Additionally, the variable expressivity observed in carriers of the same variant may be explained by allelic expression bias through autosomal random monoallelic expression (aRME) (Reinius and Sandberg 2015), a phenomenon that is thought to be common among carriers of genetic defects associated with inborn errors of immunity (IEIs). Indeed, these conditions often exhibit non-Mendelian segregation patterns and variable clinical features (Stewart et al., 2025). Moreover, the PLCγ1 isozyme is an integral component of multiple signaling pathways, and the consequences of its dysregulation are likely to be context dependent. It is likely that different *PLCG1* variants impact distinct cellular processes across various tissues and cell types, resulting in a spectrum of pathological changes. In summary, the symptoms observed in affected individuals appear to correlate, to some extent, with the severity of the variants as indicated by fly assays. However, the penetrance and expressivity of these phenotypes will require further investigation to better understand the genotype-phenotype associations of *PLCG1* variants.

## Data Availability

All data produced in the present study are available upon reasonable request to the authors.

## Data and code availability

This study did not generate datasets. All reagents developed in this study are available upon request.

## Acknowledgments

We thank the individuals and families for their participation in this study. We thank Ms. Hongling Pan for helping in the generation of transgenic fly lines. We thank Dr. Meisheng Ma for suggestions about protein structure interpretation. We thank Dr. Zhandong Liu for providing computational resources for performing the molecular dynamics simulations. We thank the Bloomington Drosophila Stock Center (BDSC) for providing stocks.

## Author contributions

Conceptualization: M.M., Y.Z., S.R.L., I.A.G., P.P., J.R., S.M., H.J.B.; Data curation: M.M., Y.Z., S.R.L., I.A.G., E.B., P.P., S.M., H.J.B.; Formal analysis: M.M., Y.Z., S.L., J.A.R., A.A., M.D., X.L., P.B.; Funding acquisition: UDN, H.J.B.; Investigation: M.M., Y.Z., S.L., M.D., D.L., M.E.; Resources: M.M., Y.Z., D.L., K.W.C., B.L.C., B.L.S., M.G., M.B., S.Y., M.F.W., S.R.L., I.A.G., P.P., J.R., S.M., H.J.B.; Supervision: H.J.B.; Visualization: M.M., Y.Z., M.D., S.L., X.P. L.D.; Writing-original draft: M.M., Y.Z.; Writing-review & editing: M.M., Y.Z., M.D., X.L., X.P., S.L., D.L., D.D., J.A.R., J.C., W-L.C., E.B., S.R.L., I.A.G., P.P., S.M., H.J.B..

## The Undiagnosed Diseases Network Consortia (Version 3.31.25)

Alyssa A. Tran, Arjun Tarakad, Ashok Balasubramanyam, Brendan H. Lee, Carlos A. Bacino, Daryl A. Scott, Elaine Seto, Gary D. Clark, Hongzheng Dai, Hsiao-Tuan Chao, Ivan Chinn, James P. Orengo, Jennifer E. Posey, Jill A. Rosenfeld, Kim Worley, Lindsay C. Burrage, Lisa T. Emrick, Lorraine Potocki, Monika Weisz Hubshman, Richard A. Lewis, Ronit Marom, Seema R. Lalani, Shamika Ketkar, Tiphanie P. Vogel, William J. Craigen, Jared Sninsky, Lauren Blieden, Sandesh Nagamani, Hugo J. Bellen, Michael F. Wangler, Oguz Kanca, Shinya Yamamoto, Christine M. Eng, Patricia A. Ward, Pengfei Liu, Adeline Vanderver, Cara Skraban, Edward Behrens, Gonench Kilich, Kathleen Sullivan, Kelly Hassey, Ramakrishnan Rajagopalan, Rebecca Ganetzky, Vishnu Cuddapah, Anna Raper, Daniel J. Rader, Giorgio Sirugo, Vaidehi Jobanputra, Allyn McConkie-Rosell, Kelly Schoch, Mohamad Mikati, Nicole M. Walley, Rebecca C. Spillmann, Vandana Shashi, Alan H. Beggs, Calum A. MacRae, David A. Sweetser, Deepak A. Rao, Edwin K. Silverman, Elizabeth L. Fieg, Frances High, Gerard T. Berry, Ingrid A. Holm, J. Carl Pallais, Joan M. Stoler, Joseph Loscalzo, Lance H. Rodan, Laurel A. Cobban, Lauren C. Briere, Matthew Coggins, Melissa Walker, Richard L. Maas, Susan Korrick, Jessica Douglas, Cecilia Esteves, Emily Glanton, Isaac S. Kohane, Kimberly LeBlanc, Rachel Mahoney, Shamil R. Sunyaev, Shilpa N. Kobren, Brett H. Graham, Erin Conboy, Francesco Vetrini, Kayla M. Treat, Khurram Liaqat, Lili Mantcheva, Stephanie M. Ware, Breanna Mitchell, Brendan C. Lanpher, Devin Oglesbee, Eric Klee, Filippo Pinto e Vairo, Ian R. Lanza, Kahlen Darr, Lindsay Mulvihill, Lisa Schimmenti, Queenie Tan, Surendra Dasari, Abdul Elkadri, Brett Bordini, Donald Basel, James Verbsky, Julie McCarrier, Michael Muriello, Michael Zimmermann, Adriana Rebelo, Carson A. Smith, Deborah Barbouth, Guney Bademci, Joanna M. Gonzalez, Kumarie Latchman, LéShon Peart, Mustafa Tekin, Nicholas Borja, Stephan Zuchner, Stephanie Bivona, Willa Thorson, Herman Taylor, Rakale C. Quarells, Ayuko Iverson, Bruce Gelb, Charlotte Cunningham-Rundles, Eric Gayle, Joanna Jen, Louise Bier, Mafalda Barbosa, Manisha Balwani, Mariya Shadrina, Rachel Evard, Saskia Shuman, Susan Shin, Andrea Gropman, Barbara N. Pusey Swerdzewski, Camilo Toro, Colleen E. Wahl, Donna Novacic, Ellen F. Macnamara, John J. Mulvihill, Maria T. Acosta, Precilla D’Souza, Valerie V. Maduro, Ben Afzali, Ben Solomon, Cynthia J. Tifft, David R. Adams, Elizabeth A. Burke, Francis Rossignol, Heidi Wood, Jiayu Fu, Joie Davis, Leoyklang Petcharet, Lynne A. Wolfe, Margaret Delgado, Marie Morimoto, Marla Sabaii, MayChristine V. Malicdan, Neil Hanchard, Orpa Jean-Marie, Wendy Introne, William A. Gahl, Yan Huang, Andrew Stergachis, Danny Miller, Elisabeth Rosenthal, Elizabeth Blue, Elsa Balton, Emily Shelkowitz, Eric Allenspach, Fuki M. Hisama, Gail P. Jarvik, Ghayda Mirzaa, Ian Glass, Kathleen A. Leppig, Katrina Dipple, Mark Wener, Martha Horike-Pyne, Michael Bamshad, Peter Byers, Runjun Kumar, Seth Perlman, Sirisak Chanprasert, Virginia Sybert, Wendy Raskind, Nitsuh K. Dargie, Chun-Hung Chan, Dr. Francisco Bustos velasq, Isum Ward, Jason Schend, Jennifer Morgan, Megan Bell, Miranda Leitheiser, Mohamad Saifeddine, Paul Berger, Rachel Li, Taylor Beagle, Alexander Miller, Beatriz Anguiano, Beth A. Martin, Brianna Tucker, Chloe M. Reuter, Devon Bonner, Elijah Kravets, Hector Rodrigo Mendez, Holly K. Tabor, Jacinda B. Sampson, Jason Hom, Jennefer N. Kohler, Jennifer Schymick, John E. Gorzynski, Jonathan A. Bernstein, Kevin S. Smith, Laura Keehan, Laurens Wiel, Matthew T. Wheeler, Meghan C. Halley, Mia Levanto, Page C. Goddard, Paul G. Fisher, Rachel A. Ungar, Raquel L. Alvarez, Sara Emami, Shruti Marwaha, Stephen B Montgomery, Suha Bachir, Tanner D Jensen, Taylor Maurer, Terra R. Coakley, Euan A. Ashley, Ali Al-Beshri, Anna Hurst, Brandon M Wilk, Bruce Korf, Elizabeth A Worthey, Kaitlin Callaway, Martin Rodriguez, Tammi Skelton, Tarun KK Mamidi, Andrew B. Crouse, Jordan Whitlock, Mariko Nakano-Okuno, Matthew Might, William E. Byrd, Albert R. La Spada, Changrui Xiao, Elizabeth C. Chao, Eric Vilain, Jose Abdenur, Kirsten Blanco, Maija-Rikka Steenari, Rebekah Barrick, Richard Chang, Sanaz Attaripour, Suzanne Sandmeyer, Tahseen Mozaffar, Alden Huang, Andres Vargas, Bianca E. Russell, Brent L. Fogel, Esteban C. Dell’Angelica, George Carvalho, Julian A. Martínez-Agosto, Layal F. Abi Farraj, Manish J. Butte, Martin G. Martin, Naghmeh Dorrani, Neil H. Parker, Rosario I. Corona, Stanley F. Nelson, Yigit Karasozen, Aaron Quinlan, Alistair Ward, Ashley Andrews, Corrine K. Welt, Dave Viskochil, Erin E. Baldwin, John Carey, Justin Alvey, Laura Pace, Lorenzo Botto, Nicola Longo, Paolo Moretti, Rebecca Overbury, Russell Butterfield, Steven Boyden, Thomas J. Nicholas, Matt Velinder, Gabor Marth, Pinar Bayrak-Toydemir, Rong Mao, Monte Westerfield, Brian Corner, John A. Phillips III, Kimberly Ezell, Lynette Rives, Rizwan Hamid, Serena Neumann, Ashley McMinn, Joy D. Cogan, Thomas Cassini, Alex Paul, Dana Kiley, Daniel Wegner, Erin McRoy, Jennifer Wambach, Kathy Sisco, Patricia Dickson, F. Sessions Cole, Dustin Baldridge, Jimann Shin, Lilianna Solnica-Krezel, Stephen C. Pak, Timothy Schedl, Allen Bale, Carol Oladele, Caroline Hendry, Emily Wang, Hua Xu, Hui Zhang, Lauren Jeffries, María José Ortuño Romero, Mark Gerstein, Michele Spencer-Manzon, Monkol Lek, Nada Derar, Odelya Kaufman, Shrikant Mane, Teodoro Jerves Serrano, Vasilis Vasiliou, Winston Halstead, Yong-Hui Jiang

