## Supplemental Figures, Methods and Materials for "Heterozygous variants in *PLCG1* affect hearing, vision, cardiac, and immune function"

**Supplemental Data**

**Additional candidate variants identified in the affected Individuals**

Individual 1 carries an intragenic duplication in *PSD3*. *PSD3* has not been associated with a Mendelian disorder but is potentially associated with an autosomal dominant arthrogryposis ^1^. Hence, it may underlie the joint defects observed in proband 1.

Individual 2 has compound heterozygous missense variants in *ERAP2* and *SEMA3G*. *ERAP2* [MIM: 609497] has not been associated with a Mendelian disorder. It encodes an ER-residential metalloaminopeptidase that functions in the major histocompatibility class I antigen presentation pathway. Some variants in *ERAP2* are associated with a susceptibility to autoimmune diseases such as ankylosing spondylitis and Crohn’s disease ^2-4^. Given that proband 2 exhibits neuroinflammation and encephalitis, these phenotypes may be associated with the *ERAP2* variants. *SEMA3G* (Semaphorin 3G) has not been associated with a Mendelian disorder. However, a homozygous missense variant in *SEMAG3* was observed in two affected siblings from a consanguineous family. The siblings exhibited dysmorphic features as well as developmental delay ^5^.

Individual 3 carries a *de novo* missense variant in *PKP2* [MIM: 602861]. *PKP2* encodes Pakophilin-2 and has been associated with dominant arrhythmogenic right ventricular dysplasia 9 [MIM: 609040] ^6-8^. However, this proband was born with septal defects.

No additional candidate variants were reported for Individuals 4-7.

**Supplemental Figures and Legends**

**Figure S1.** ***s******l^T2A^* is a loss-of-function allele causing wing and eye phenotypes**

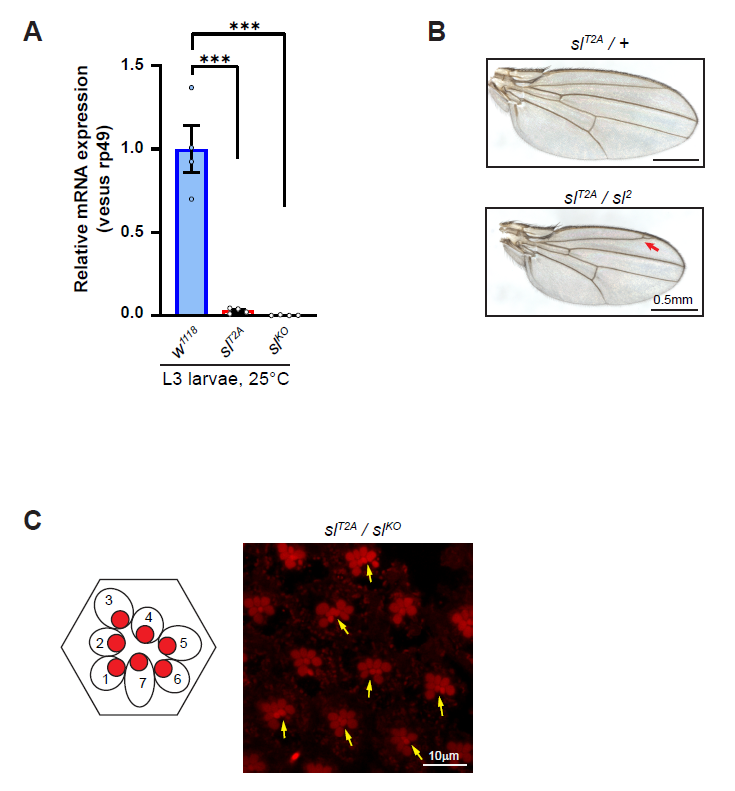

(A) Relative *sl* mRNA expression levels are <5% and <1% in *sl**^T2A^* and *sl**^KO^* mutant larvae when compared to controls (*w^1118^*). The primers used for real-time PCR are shown in Figure 1C. Each dot represents a replicate per genotype. Unpaired t test, ***p<0.001, mean ± SEM.

(B) Representative images showing that *sl^T2A^*/*sl^2^* trans-heterozygous mutant flies have smaller wing and ectopic veins (indicated by arrow). Scale bars, 0.5mm.

(C) Representative images showing that *sl^T2A^*/*sl^KO^* trans-heterozygous mutant flies have extra photoreceptors (indicated by arrows). The schematic of the section of an ommatidia presenting seven photoreceptors (the R8 photoreceptor is not visible in such section) is shown. The photoreceptor rhabdomeres stain positive for phalloidin labeling F-actin. Scale bar, 10μm.

**Figure S2. Proband-associated variants exhibit deleterious impacts in adults**

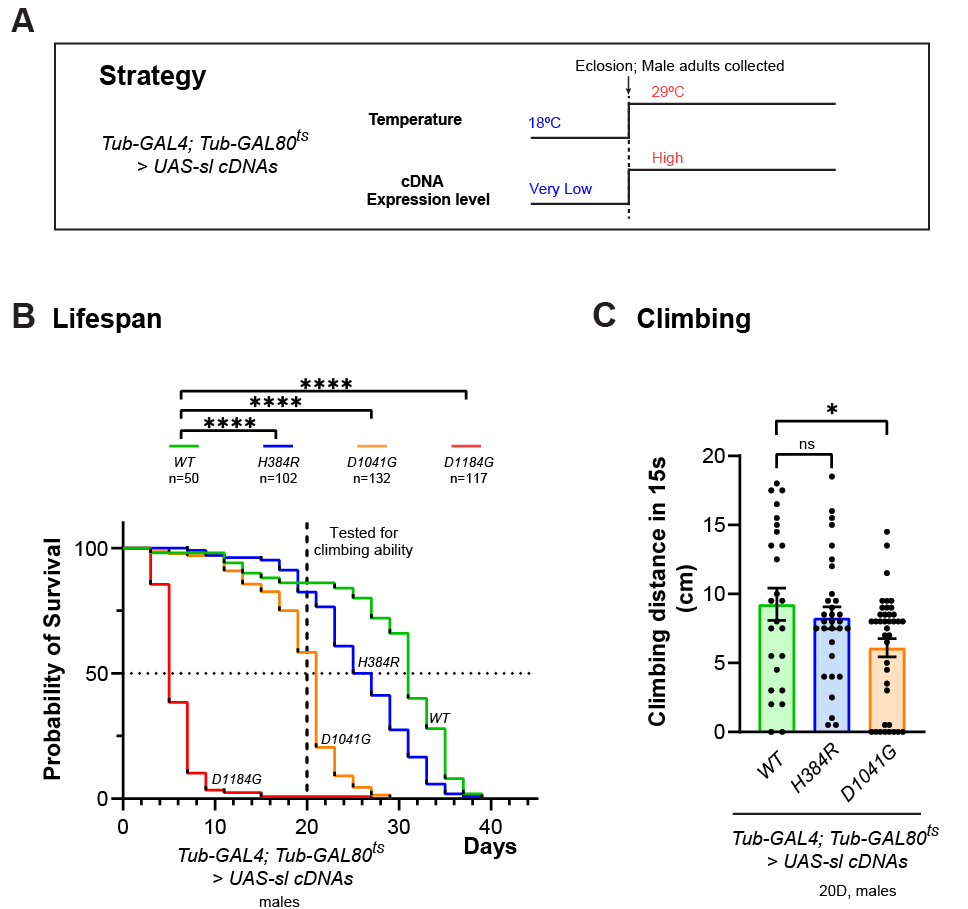

(A) Cross strategy of the temperature shifting assay to express *UAS-sl* cDNAs only in the adult stages. GAL80^ts^ is a temperature-sensitive inhibitor of GAL4, which allows temporal control of *UAS-* transgene expression by temperature manipulation. *GAL80^ts^* represses GAL4 activity at the permissive temperature (18°C) but becomes inactive at restrictive temperature (29°C), thereby allowing GAL4 to activate expression of *UAS-transgenes*. In this assay, *Tub-GAL4, Tub-GAL80^ts^* driver was used to express wild-type or variant *sl cDNAs* specifically in adult flies by shifting the temperature after eclosion.

(B) Adult-stage expression of *sl* variants induces lifespan reduction. The adult flies with expression of *sl^D1184G^* mostly die within one week after eclosion. The flies expressing *sl^D1041G^* or *sl^H384R^* have median lifespans of approximately 21 days and 26 days, respectively, both shorter than those expressing *sl^WT^*, which have a median lifespan of 31 days. The sample size for each genotype is indicated (n). Longrank test, ****p<0.0001.

(C) Adult-stage expression of *sl^D1041G^* induces some locomotion defect. Flies aged to 20 days were assessed for locomotor ability using a climbing assay. Expression of *sl^H384R^* did not induce significant difference compared to *sl^WT^*. We did not test *sl^D1184G^* as the flies are short lived. Each dot represents the measurement of one fly. Unpaired t test, *p<0.05, ns: not significant, mean ± SEM.

These results suggest that the two strong variants (*sl^D1041G^* and *sl^D1184G^*) contribute to both developmental and acute effects while the *sl^H384R^* variant mainly contributes to developmental stages.

**Figure S3. The toxicity of human *PLCG1* cDNAs in flies correlates with expression levels (related to Figure 4)**

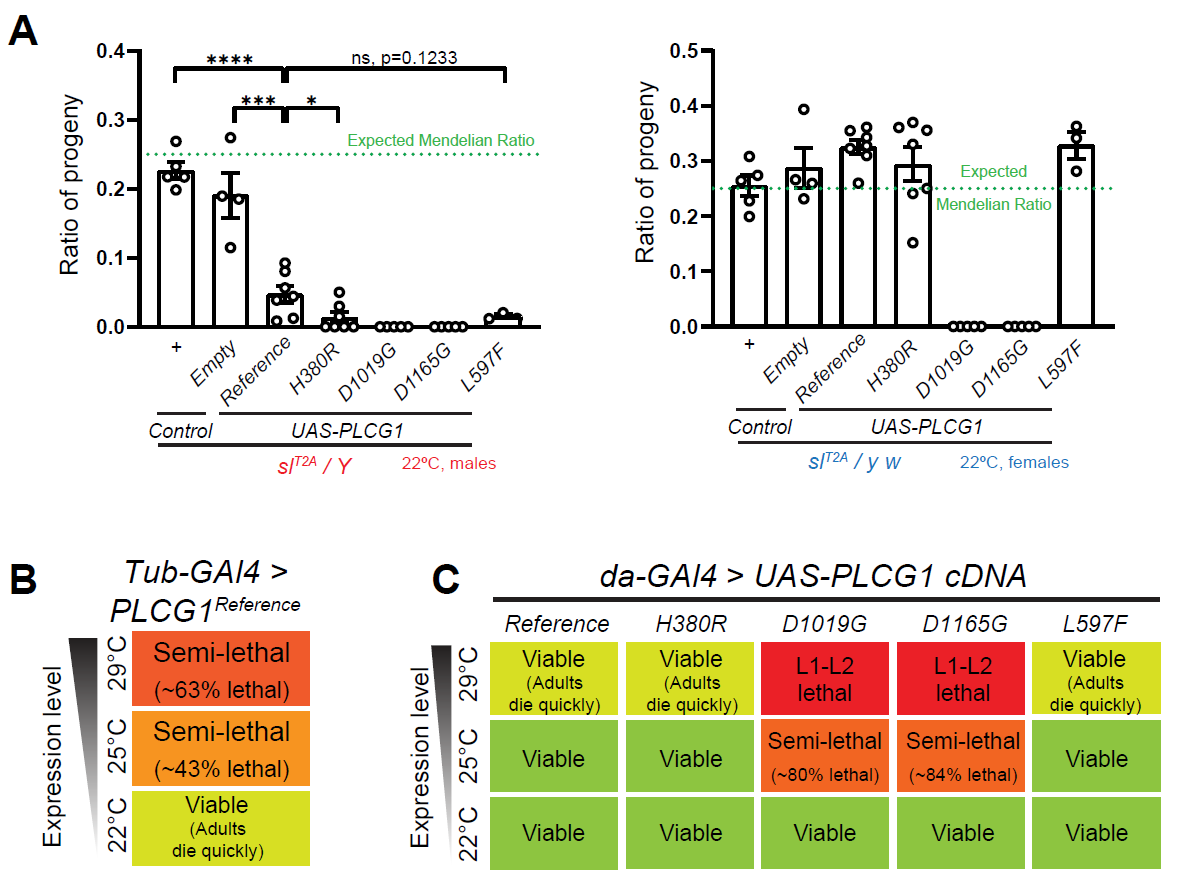

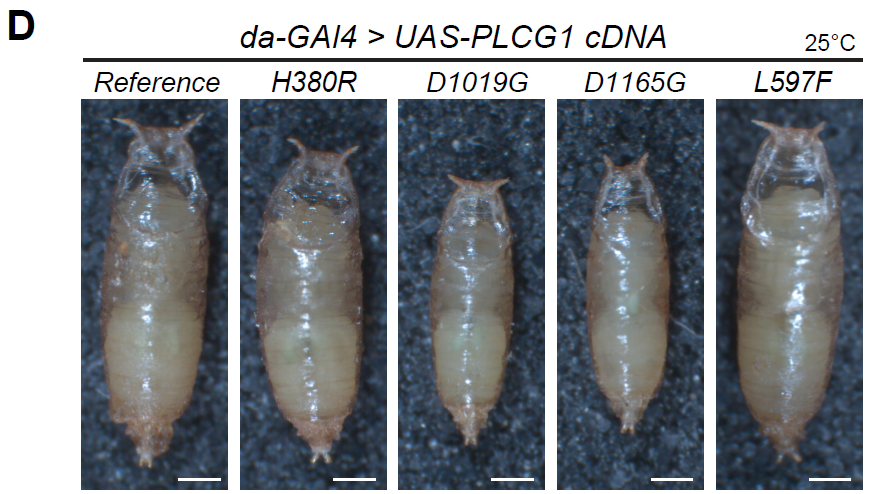

(A) Summary of the eclosion rate of animals expressing *sl* cDNAs in *sl^T2A^* mutant (left panel) or heterozygous (right panel) flies at 22°C. The phenotypes are similar to, but slightly milder compared to the same assays performed at 25°C (Figure 4B). Expression of *PLCG1^D1019G^* or *PLCG1^D1165G^* caused lethality in both conditions, whereas expression of *PLCG1^Reference^*, *PLCG1^H380R^*, or *PLCG1^L597F^* led to reduced eclosion rate in mutant hemizygous but not in heterozygous flies. Each dot represents one independent replicate. Unpaired t test, ****p < 0.0001, ***p < 0.001, *p<0.05, ns: not significant, mean ± SEM.

(B) Summary of the viability of expressing *PLCG1^Reference^* ubiquitously using *Tub-GAL4*. Expression levels of the *UAS- PLCG1^Reference^* transgenes can be manipulated at different temperatures.

(C) Summary of the viability of expressing *PLCG1* reference and variant cDNAs using a weak ubiquitous driver *da-GAL4*. Expression levels of the *UAS- PLCG1^Reference^* transgenes can be manipulated at different temperatures.

(D) Representative images of the pupae of *da-GAL4 > UAS-PLCG1 cDNAs* flies at 25°C. Expression of *PLCG1^D1019G^* or *PLCG1^D1165G^* caused reduced pupal size.

**Figure S4. Expressing human *PLCG1* does not rescue wing or eye phenotypes associated with *sl^T2A^* (related to Figure 4)**

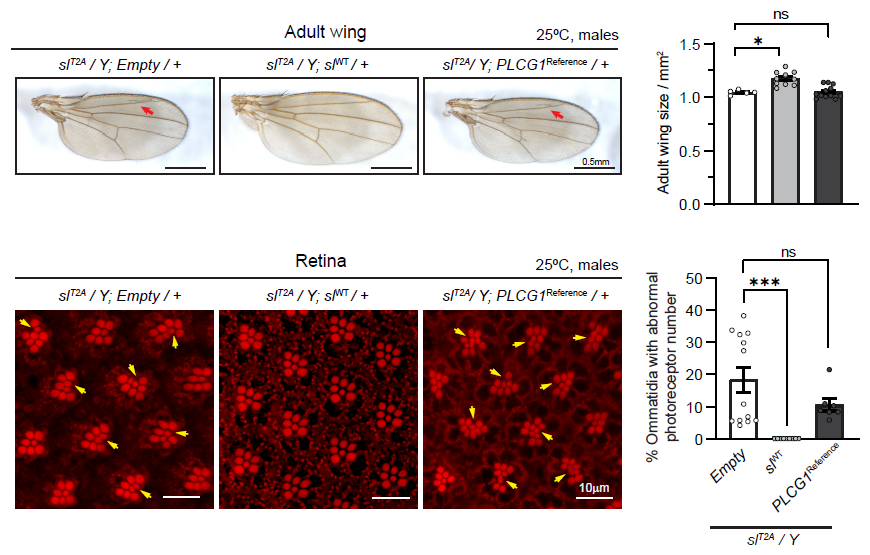

Representative images showing the adult wings (upper panel) and the photoreceptors (lower panel) expressing *PLCG1^Reference^* or *sl^WT^*. Expression of *sl^WT^* rescues the loss-of-function phenotypes including wing size reduction, ectopic veins (indicated by red arrows) and extra photoreceptors (indicated by yellow arrows), whereas expression of the *PLCG1^Reference^* or *UAS-Empty* shows no rescue. Scale bars, 0.5mm for the wing images and 10μm for the photoreceptor images. The photoreceptor rhabdomeres stain positive for phalloidin labeling F-actin. Quantification of the wing size (upper right panel) and the photoreceptors (lower right panel) are shown. Each dot represents measurement of one wing or retina sample, respectively. Unpaired t test, ***p<0.001, *p<0.05. ns: not significant.

**Notes for Figure S3 and S4:**

We assessed if human *PLCG1* could effectively serve as a functional substitute for fly *sl* and rescue the loss-of-function phenotypes observed in *sl* mutant flies. However, as shown in Figure 4B, only a small fraction of the *sl^T2A^/Y* mutant hemizygotes expressing *PLCG1^Reference^* can survive to adults, and the escapers die within one week. Since overexpression of fly *sl^WT^* does not cause viability issues (Figure 4A), the reduced viability associated with *PLCG1^Reference^* in *sl^T2A^* mutant male progeny may be due to elevated expression levels of the human proteins.

To assess the “high expression level toxicity” hypothesis, we raised the flies at different temperatures and tested with various GAL4 driver. The GAL4-UAS system is highly temperature-dependent since the promoter in the UAS construct contains an Hsp-70 promoter (Fischer et al. 1988), and the expression levels increase with higher temperatures and decrease with lower temperatures (Nagarkar-Jaiswal et al. 2015). We assessed the viability of the progeny with expression of *PLCG1* or *sl* cDNA in hemizygous mutant flies (*sl^T2A^/Y>UAS-cDNAs*) at 22°C, the survival ratio was increased but the increase was subtle (Figure S3A). To avoid the dosage compensation effect that alters the expression level of *sl^T2A^* between males or females, we used a ubiquitous GAL4 driver, *Tub-GAL4*, to ectopically drive the expression of *PLCG1^Reference^* ubiquitously at different temperatures. As shown in Figure S3B, the *Tub-GAL4>UAS-PLCG1^Reference^* flies exhibited a high lethality ratio when raised at 29°C (~63% lethal) and the surviving flies die within one week. This lethality ratio decreased to ~43% when the flies were raised at 25°C whereas >90% of the flies were able to eclose as adults at 22°C. This shows that the toxicity is highly dependent on protein levels. Indeed, the expression levels can be further lowered by using a weak ubiquitous GAL4 driver, *da-GAL4*. The *da-GAL4* > *UAS-PLCG1^Reference^* flies were viable at the three tested temperatures (29°C, 25°C, and 22°C), but the enclosed adults at 29°C mostly died within one week. *da-GAL4 >* *UAS-PLCG1^H380R^* or *UAS-PLCG1^L597F^* exhibited similar phenotypes. In contrast, *da-GAL4 >* *UAS-PLCG1^D1019G^* or *UAS-PLCG1^D1165G^* flies were lethal at 29°C, semi-lethal (~80%-85% lethal) at 25°C, and viable at 22°C (Figure S3C). The animals that escape lethality at 25°C have smaller pupae (Figure S3D) and a reduced adult body size, arguing that growth is impeded. In summary, these data support the hypothesis that elevated expression levels of the human *PLCG1* is toxic in flies.

In addition to the toxicity, *PLCG1^Reference^* fails to rescue the phenotypes observed in the wings or eyes of the *sl* mutant flies (Figure S4), which is fully rescued by fly *sl^WT^* (Figure 2). Expression of the *PLCG2* cDNA in *sl^T2A^* mutant hemizygous males exhibited similar toxicity (Figure 4B) and limited ability to rescue the phenotypes caused by loss of *sl*. This suggests that despite their high DIOPT scores, the two human genes encoding PLCγ isozymes cannot fully substitute for the fly PLCγ ortholog. It is possible that during the course of evolution, *PLCG1* has acquired more specialized functions. For example, an essential step for enzymatic activation of mammalian PLCγ is the binding of its nSH2 domain to specific phosphotyrosines on the RTKs through a specific binding motif (Songyang et al. 1993). However, Thackeray *et al.* found that the consensus motif is absent in the intracellular domain of the *Drosophila* EGF receptor homolog DER (Thackeray et al. 1998), one of the three RTKs in *Drosophila* which is required for wing vein differentiation and photoreceptor formation (Dickson and Hafen 1994; Freeman 1996; Schweitzer and Shilo 1997). Nevertheless, expression of the *PLCG1* variant cDNAs leads to more severely reduced eclosion rate compared to expression of the reference cDNA (Figure 4B), suggesting that they are detrimental variants.

**Figure S5. Intracellular Ca^2+^ reporter assay suggests that the p.(Asp1019Gly) and p.(Asp1165Gly) are hyperactive variants (related to Figure 5)**

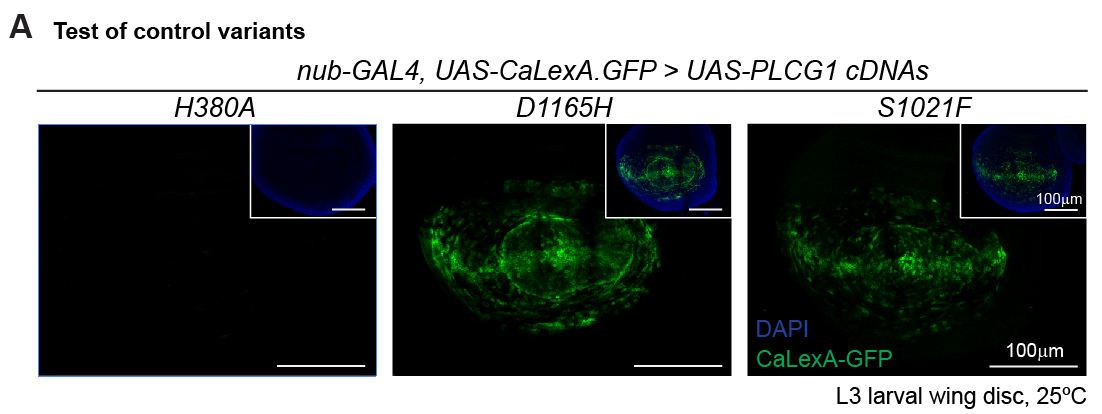

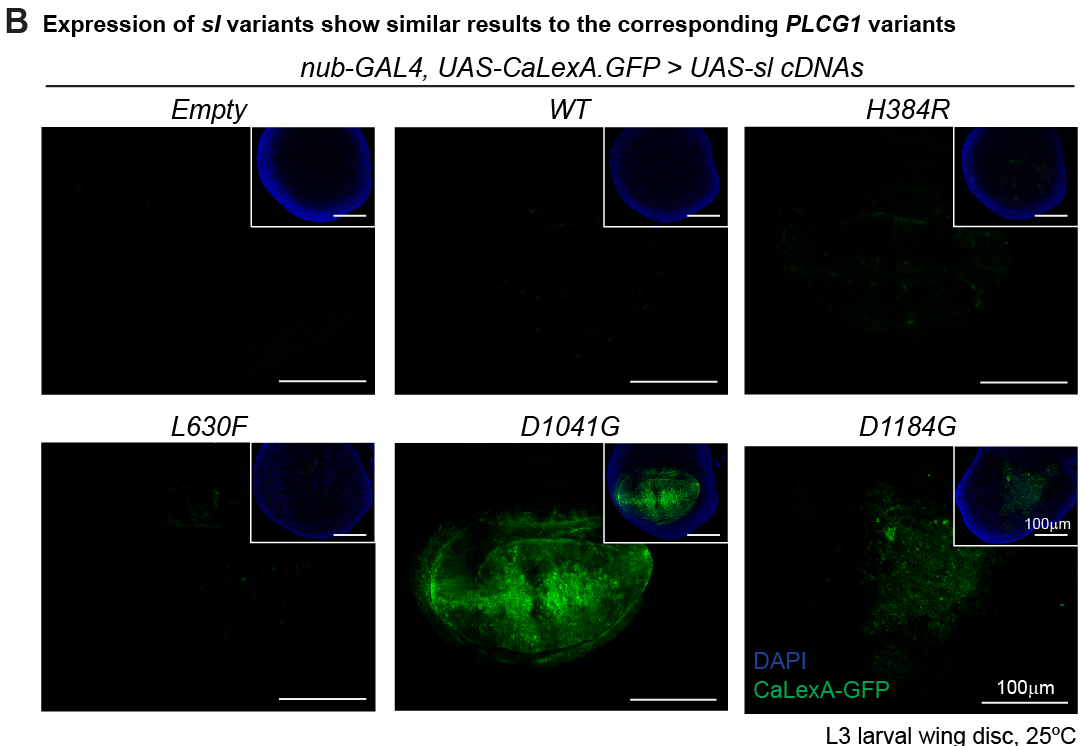

(A) Three previously-reported *PLCG1* variants (*PLCG1^H380A^*, *PLCG1^D1165H^* and *PLCG1^S1021F^*) were tested as controls. The Ca^2+^ reporter *CaLexA.GFP* was co-expressed with *PLCG1* cDNAs in the wing disc pouch region. Expression of *PLCG1^D1165H^* or *PLCG1^S1021F^* caused elevated CaLexA.GFP signal (green), whereas expression of *PLCG1^H380A^* did not. Nuclei were labeled with DAPI (blue). Insets show merged images of DAPI and GFP channels. Scale bars, 100μm.

(B) The fly variants were analyzed using the *CaLexA.GFP* reporter*.* Expression of *sl^D1041G^* or *sl^D1184G^* caused elevated CaLexA.GFP signal (green), similar to the corresponding *PLCG1* variants. Note that the wing discs expressing *sl^D1184G^* are morphologically aberrant and have a diminished GFP signal. Nuclei were labeled with DAPI (blue). Insets show merged images of DAPI and GFP channels. Scale bars, 100μm.

**Figure S6. Wing and eye phenotypes associated with ectopic expression of *PLCG1* variants (related to Figure 5)**

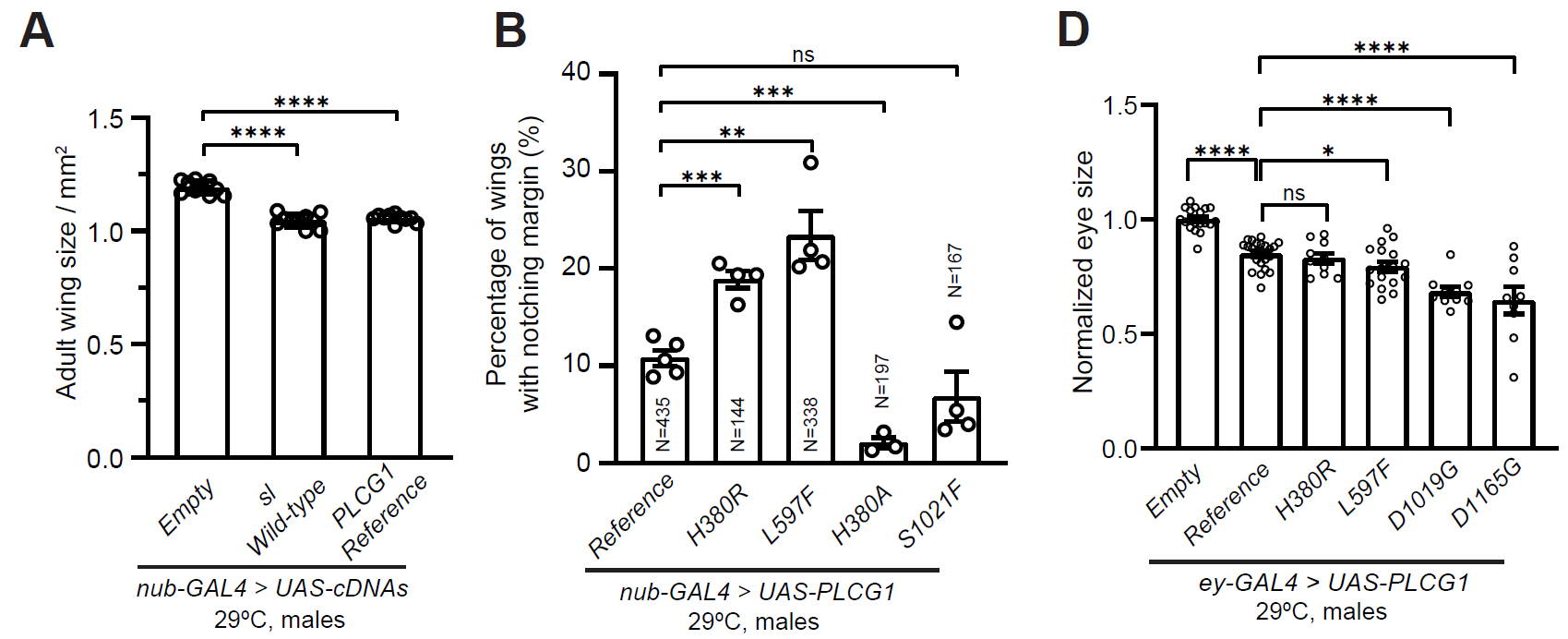

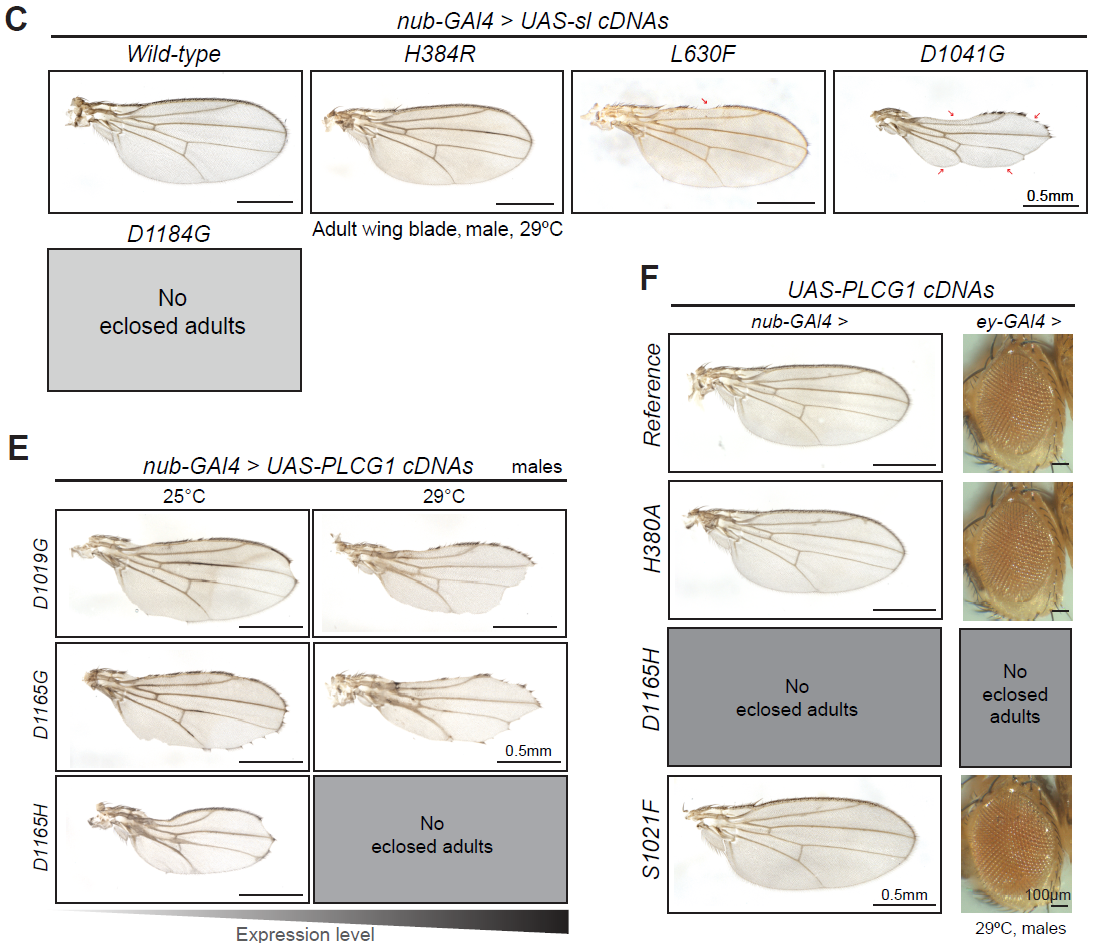

(A) Quantification of the wing blade size in non-notched wings from the samples overexpressing *PLCG1^Reference^* or *sl^WT^*. Wing-specific expression of *PLCG1^Reference^* or *sl^WT^* caused an approximate 15% wing size reduction compared to the *UAS-Empty* control construct. Each dot represents one measured adult wing. Unpaired t test, ****p < 0.0001, mean ± SEM. Scale bars, 0.5mm.

(B) Quantification of the percentage of adult wings with margin notching phenotype. Wing-specific expression of *PLCG1^Reference^* caused ~10% wings to display a notched margin. Expression of *PLCG1^H380R^* or *PLCG1^L597F^* caused approximately 18% and 23% of the wings to display notched margins, respectively. Expression of the enzymatic-dead *PLCG1^H380A^* caused less than 10% wings with notched margin, whereas expression of the hyperactive *PLCG1^S1021F^* is not significantly different from *PLCG1^Reference^*. Each dot represents one independent replicate. The total number of flies (N) was counted per genotype. Unpaired t test, ***p < 0.001, **p < 0.01, ns: not significant, mean ± SEM.

(C) Representative images of adult wing blades showing morphological phenotypes caused by wing-specific expression of *sl* cDNAs. Expression of *sl^D1041G^* caused severe wing defects including notched margins (arrows) and fused/thickened veins (arrowheads). Expression of *sl^H384R^* or *sl^L630F^* exhibited partial penetrance. Expression of *sl^D1184G^* caused lethality before eclosion. Scale bars, 0.5mm.

(D) Quantification of eyes size with expression of *PLCG1* cDNAs. Eye size were normalized to the size of the *ey-GAL4>UAS-Empty* control. Each dot represents the measurement of one adult eye. Unpaired t test, ****p < 0.0001, *p < 0.05, ns: not significant, mean ± SEM.

(E) Representative images showing that wing-specific expression of *PLCG1^D1165H^* caused similar but more severe morphological phenotypes compared to *PLCG1^D1019G^* or *PLCG1^D1165G^*. Expression levels of *UAS-cDNA* can be manipulated by raising the animals at different temperatures. Scale bars, 0.5mm.

(F) Representative images showing the phenotypes associated with ectopic expression of the control *PLCG1* variants. Expression of *PLCG1^H380A^* or *PLCG1^S1021F^* did not induce obvious morphological phenotypes in adult wings or eyes compared to *PLCG1^Reference^*, whereas no eclosed adults were collected when overexpressing *PLCG1^D1165H^* at 29°C in these contexts. Scale bars, 0.5mm for wing images, 100μm for eye images.

**Notes for Figure S5 and S6:**

To better understand whether the phenotypes caused by the variants in fly models are associated with the enzymatic activity of the PLCγ1 isozyme, we characterized the phenotypes of transgenic flies expressing control constructs: *PLCG1^H380A^* (enzymatic dead), *PLCG1^D1165H^* (hyperactive) and *PLCG1^S1021F^* (hyperactive). Ectopic expression of the enzymatic-dead *PLCG1^H380A^* or hyperactive *PLCG1^S1021F^* in wings or eyes did not cause obvious morphological abnormalities, whereas expression of the hyperactive *PLCG1^D1165H^* caused very severe phenotypes (Figure S6F). Overexpression of *PLCG1^D1165H^* in the eyes or wings causes lethality at 29°C, arguing that it is highly toxic. These flies survive when they are raised at 25°C, yet the wings show severe morphological defects, including notched wing margins, thickened veins as well as reduced wing size (Figure S6E). These phenotypes are similar to, but more severe than, the defects observed in the wings expressing *PLCG1^D1019G^* or *PLCG1^D1165G^* (Figure 5B). Notably, expression of *PLCG1^H380R^* or *PLCG1^L597F^*, which did not exhibit a hyperactive effect in the *CaLexA* reporter assay compared to *PLCG1^S1021F^* (Figure 5A and S5A), caused a partially penetrant wing notching phenotype i (Figure 5B and S6B). Additionally, *PLCG1^L597F^* expression led to a reduction in eye size (Figure 5C and S6D). These observations suggest that the morphological phenotypes in wings and eyes are not directly correlated with the enzymatic activity of the PLCγ1 isozyme, but may instead be associated with neomorphic effects. Similarly, the increased lethality observed in *sl^T2A^*-driven expression of cDNAs may also be associated with neomorphic effect. As shown in Figure 4B, when driven by *sl^T2A^*, *PLCG1^H380A^* led to reduced viability in hemizygous mutant flies but not in heterozygous ones, whereas *PLCG1^D1165H^* caused 100% lethality in both genotypes. However, the hyperactive *PLCG1^S1021F^* did not show distinguishable phenotypes compared to *PLCG1^Reference^* in this assay. In summary, the *PLCG1* variants identified in this study exhibit neomorphic effects with variable degrees of severity, whereas the p.(Asp1019Gly) and p.(Asp1165Gly) variants are hyperactive and also cause neomorphic phenotypes.

**Figure S7. *In silico* analyses of *PLCG1* variants**

**A. Molecular dynamics simulations of the p.(Asp1019Gly) and p.(Asp1165Gly) variants**

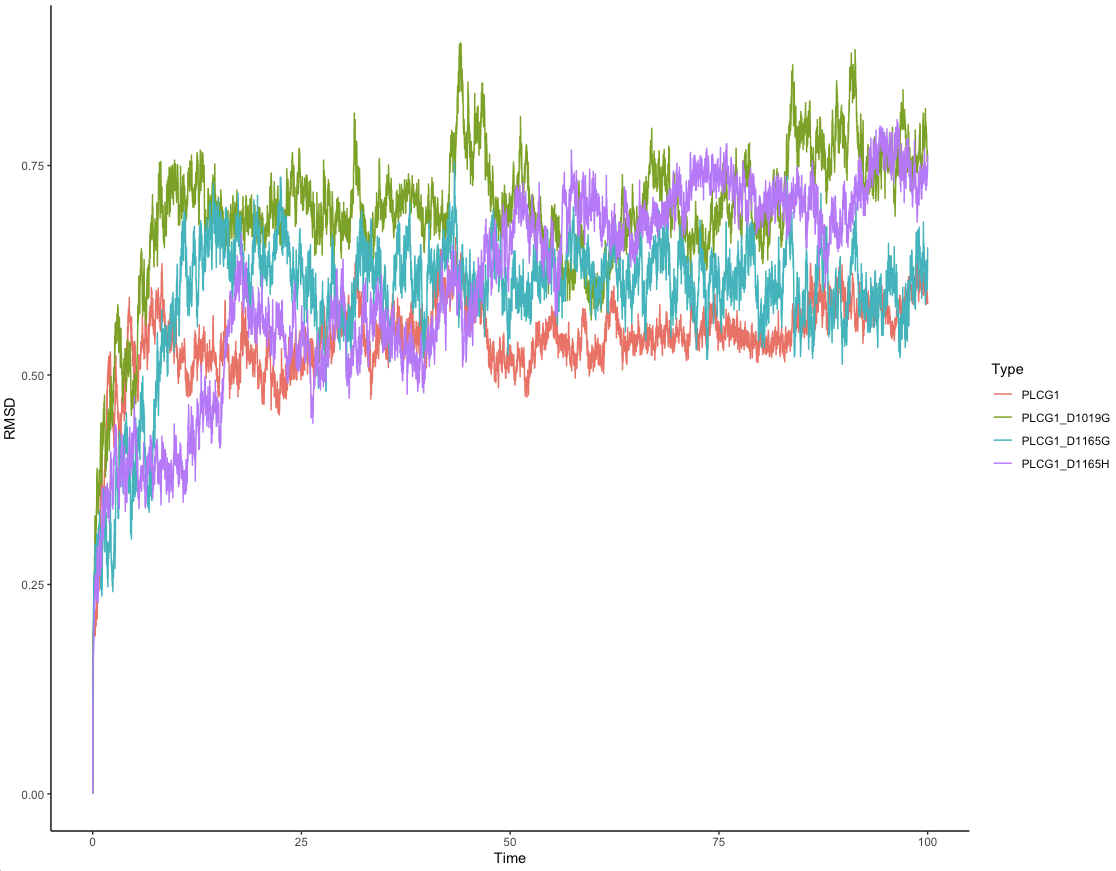

**B. Protein stability and folding prediction utilizing DDMut**

| **chain** | **aa_from** | **position** | **aa_to** | **Predicted Stability Change (ΔΔG^Stability wt->mt^) *kcal/mol*** |
| --- | --- | --- | --- | --- |
| A | H | 380 | R | -0.22 |
| A | D | 1019 | G | -0.91 |
| A | D | 1165 | G | -1.54 |
| A | L | 597 | F | -1.26 |
| A | H | 380 | A | -0.13 |
| A | D | 1165 | H | -1.59 |
| A | S | 1021 | F | 0.14 |

*The wild-type Rat Plcg1 protein structure (PDB: 6PBC) was used as the reference protein.

(A) Molecular dynamics simulations for the p.(Asp1019Gly) and p.(Asp1165Gly) variants. These variants exhibit increased disorganization with a higher root mean square deviations (RMSD) compared to the reference PLCG, similar to the hyperactive p.(Asp1165His) variant. Color code: PLCG1 reference - red; p.(Asp1019Gly) - green; p.(Asp1165Gly) - cyan; p.(Asp1165His) - purple.

(B) Prediction of the effects of PLCG1 variants on protein stability and folding using DDMut. DDMut is a platform designed to predict protein stability based on the Gibbs Free Energy change (ΔΔG), and larger ΔΔG indicates a more pronounced impact on protein stability. The protein structure of rat Plcg1 [Protein Data Bank (PDB) ID 6PBC] was used as reference. The p.(Asp1019Gly), p.(Asp1165Gly), and p.(Asp1165His) variants are predicted to induce destabilizing, with ΔΔG^Stability wt->mt^ values of -0.91 kcal/mol, -1.54 kcal/mol, and -1.59 kcal/mol, respectively, suggesting impaired protein folding or altered conformational dynamics. These results are consistent with their proposed roles in disrupting autoinhibitory interactions (Hajicek et al. 2013; Siraliev-Perez et al. 2022). The ΔΔG^Stability wt->mt^ value for p.(His380Arg) and the enzymatically inactive p.(His380Arg) are predicted to be -0.22 kcal/mol and -0.13 kcal/mol, respectively. Given that the His380 residue’s influence on the phospholipase activity might not through the autoinhibition mechanism, the impact of His380 residue variants on the protein folding and stability may not elucidate their effects on protein function. The ΔΔG^Stability wt->mt^ value for p.(Leu380Phe) is predicted to be -1.26 kcal/mol, which is not inconsistent with the potential impact of this variant on interaction between nSH2 domain and RTK activators. However, the p.Ser1021Phe variant has a predicted ΔΔG^Stability wt->mt^ value of 0.14 kcal/mol, indicating a slight stabilizing effect. These analyses indicate that the variants may act through various pathogenic mechanisms, but further investigations are needed.

**Supplemental Tables**

Table S1. Pathogenicity prediction of the proband-variants

Table S2. Mammalian PLC coding genes and their fly orthologs

Table S3. Summary of fly assay phenotypes

Table S4. Fly strains used in the experiments

Table S5. Primers used in the experiments

**Material and methods**

**Recruitment of the probands**

Individuals 1 and 2 were recruited through the Undiagnosed Diseases Network (UDN) and were evaluated through the clinical research protocol of the National Institutes of Health Undiagnosed Diseases (15-HG-0130), which was approved by the National Human Genome Research Institute (NHGRI). Individuals 3-7 were recruited through GeneMatcher. Formal consents for genetic testing and publication were obtained from all individuals or their family members.

***Drosophila* husbandry and generation of transgenic flies**

The fly strains used in this study (listed in Table S3) were generated in house or obtained from the Bloomington Drosophila Stock Center (BDSC). All the flies were raised and maintained on standard fly food at room temperature unless specified. The *sl^T2A^* allele was outcrossed with *w^1118^* to clean up the genetic background. The strains used in this study were listed in Supplemental Table S3.

To generate the *UAS-cDNA* transgenic lines, human *PLCG1* cDNA was obtained from Horizon Discovery (MHS6278-213246131, clone ID 9052656), and fly *sl* cDNA was obtained from *Drosophila* Genomics Resource Center (DGRC, RE62235). The coding sequence (CDS) of *PLCG1^Reference^* and *sl^WT^* were amplified using iProof™ High-Fidelity DNA Polymerase Kit (BioRad, #1725301), purified using QIAEX II Gel Extraction Kit (QIAGEN, #20021), sub-cloned into the Gateway compatible entry vector pDONR223 by BP cloning (BP clonase II, Thermo Fisher Scientific, #11789020) and sequentially cloned into the destination vector pGW-attB-HA by LR cloning (LR clonase II, Thermo Fisher Scientific, #11791100) (Bischof et al. 2013). The variants were generated by site-directed mutagenesis strategy using Q5 Hot Start High-Fidelity 2x Master Mix (NEB, #M0494S) and *DpnI* restriction enzyme (NEB, # R0176L). All the constructs were sanger verified and injected, and inserted into the VK33 (*PBac{Yang, 2013 #228}{y[+]-attP}VK00033*) docking site using ϕC31 mediated transgenesis (Venken et al. 2006; Bischof et al. 2007). Primers are listed in Supplemental Table S4.

***Drosophila* behavioral assays**

The climbing assay to examine the negative geotaxis and locomotion ability of the flies was performed as previously described (Madabattula et al. 2015; Lu et al. 2022) with some modifications. 18-22 flies per vial were transferred to an empty plastic vial and given 20min to rest prior to being tested. The flies were tapped to the bottom of the vial and were allowed to climb for 15s. The percentage of flies per vial that climbed over 5cm were calculated. The maximum distance from the bottom to the top is 18.5cm.

For the lifespan assay, newly eclosed male flies were collected and maintained at 25°C (10 flies per vial). The flies were transferred to a new vial and the number of dead flies was counted every two days.

For the temperature-shifting related assay, flies were raised at 18°C until eclosion. Newly eclosed males were collected and maintained at 29°C for the lifespan assay and climbing assays conducted at specified ages.

**Immunostaining**

Fly tissues were dissected in 1x PBS, fixed in 4% paraformaldehyde for 20min at room temperature, and washed in PBS (3 x 10min). For antibody staining, samples were treated with PBST (Triton X-100 in PBS, 0.1% for larval tissues, 2% for adult brain), 5% normal goat serum, and incubated in primary antibody overnight at 4°C. Samples were washed with 0.1% PBST (3 x 10min) and incubated with secondary antibody for 2h at room temperature (in darkness) and washed in 0.1% PBST (3 x 10min). Primary antibodies: rat anti-*Drosophila* Elav (1:250, DSHB, #7E8A10); mouse anti-*Drosophila* Repo (1:50, DSHB, #8D12). Secondary antibodies: goat anti-rat-647 (1:250, Jackson ImmunoResearch, #112-605-003), goat anti-mouse-Cy5 (1:250, Invitrogen, #A10524). Larval discs were mounted in Vectashield (Vector Labs #H1200 and #H1000). Larval CNS and adult brain were mounted in Rapiclear (Cedarlane, #RC147001). For adult retinas, flies are reared at 25 °C under 12-h light/dark conditions. Retinas were isolated from 5-7 day old flies. Heads were dissected in PBS and fixed in 3.7% formaldehyde overnight at 4°C. The samples were rinsed with 0.1% PBST and the retinas were subsequently dissected and incubated with PBST-diluted phalloidin 647 (1:100, Invitrogen, #A22287) for 1h. Retinas were washed in 0.1% PBST and mounted in Vectashield. The images were obtained with a confocal microscope (Leica SP8X or Zeiss Airyscan LSM 880) and processed using the ImageJ-FIJI software (Schneider et al. 2012).

**Imaging of adult fly wings and eyes**

To prepare the samples of adult fly wings, the wing blades were dissected and mounted in a glycerol/ethanol 1/1 mixture. Only wings from the same gender were compared to each other since females have larger wings than males when raised in the same conditions. To prepare the samples of adult fly eyes, the flies were frozen and placed onto a double-side stick tape with one eye facing up. The samples were imaged using bright field Stereomicroscope (Leica MZ16 or Leica Z16 APO). Image Pro Plus 7.0 software was used to for extended depth-of-ﬁeld images. The image processing and the measurement of the total areas of the wing blades or eyes were conducted using the ImageJ-FIJI software (Schneider et al. 2012).

**Real-time PCR**

Real-time PCR was performed as previously described (Ravenscroft et al. 2020) with modifications. All-In-One 5X RT MasterMix (abm, #G592), iTaq Universal SYBR Green Master Mix (BioRad#1725120) and BioRad C1000 Touch Cycler were used. Primers are listed in Supplemental Table S4.

**Molecular dynamics simulations**

The three-dimensional structures of the wild-type and variant forms of the PLCG1 protein were predicted using AlphaFold3 (Abramson et al. 2024). All simulations were performed using GROMACS (Pronk et al. 2013) version 2020.6. Initial PDB files were processed to remove water molecules and hydrogen atoms. The AMBER14SB_parmbsc1 force field (Maier et al. 2015) was employed for parameterization. TIP3P water model was used to solvate the system in a cubic box with a minimum distance of 1.0 nm between the protein and box edges. Counterions (Na⁺ and Cl⁻) were added to neutralize the system’s net charge. Energy minimization was conducted using the steepest descent algorithm to eliminate unfavorable contacts. Subsequently, the system underwent equilibration in two phases: NVT Equilibration: Maintained at 300 K using the velocity-rescaling thermostat for 100 ps; NPT Equilibration: Pressure was stabilized at 1 bar using the Parrinello-Rahman barostat for 100 ps.

An unrestrained production MD simulation was carried out for 100 ns under constant temperature (300 K) and pressure (1 bar) conditions. The LINCS algorithm was used to constrain all bonds involving hydrogen atoms, allowing a time step of 2 fs. Long-range electrostatics were treated using the Particle Mesh Ewald (PME) method with a cutoff of 1.0 nm for both Coulomb and van der Waals interactions.

Post-simulation analyses included root mean square deviation (RMSD) calculations to assess structural stability and radius of gyration (Rg) to evaluate compactness. All analyses were performed using built-in GROMACS tools. The results are plotted by ggplot2 R package.
